# Do changes in antiseizure medication affect seizure timing?

**DOI:** 10.1101/2025.07.10.25331313

**Authors:** Ashley Reynolds, Rachel E. Stirling, Samuel Håkansson, Philippa Karoly, Alan Lai, David B. Grayden, Mark J. Cook, Ewan S. Nurse, Andre Peterson

**Affiliations:** Department of Biomedical Engineering, The University of Melbourne, Melbourne, Australia; Department of Medicine, The University of Melbourne, Melbourne, Australia; Department of Neurosciences, St. Vincent’s Hospital, The University of Melbourne, Melbourne, Australia; Graeme Clark Institute, The University of Melbourne, Melbourne, Australia; Seer Medical, Melbourne VIC 3000, Australia; Department of Clinical Neuroscience, Institute of Neuroscience and Physiology, Sahlgrenska Academy, Gothenburg University, Gothenburg, Sweden

## Abstract

**Key points:**

1. Antiseizure medications may change how strongly seizures synchronise with seizure cycles estimated from seizure diaries.
2. One seizure rate can be produced by different seizure cycles, suggesting a one-to-many relationship between seizure rate and cycles.
3. Using seizure cycles to time medication and assess efficacy could prove challenging with paper-based monitoring due to the complexity of cycles and potential drug effects. A seizure cycle tracking algorithm combined with an electronic seizure diary might support this task.

Evaluating effectiveness of anti-seizure medications in epilepsy often relies on seizure frequency, reported through seizure diaries before and after treatment initiation. Measuring efficacy with seizure frequency can be challenging and unreliable as seizures tend to occur in cyclical patterns— seizure cycles—making it difficult to distinguish drug effects from natural fluctuations. Incorporating cycle information could aid treatment evaluation, but antiseizure medications (ASMs) may alter seizure cycles, warranting further study.

We conducted an observational study using seizure and ASM tracking app data (Feb. 2023) from 86 individuals with epilepsy. Participants were grouped based on ASM regimen and ≥50% seizure rate reduction at 4 months after a drug change (drug-switching-responders, n=7/45; drug-switching-non-responders, n=38/45) or random timepoint (drug-sustained-responders, n=8/41; drug-sustained-non-responders, n=33/41). We compared groups on three seizure cycle variables detected via diaries: 1. how strongly seizures synchronise with a cycle, measured by the Synchronisation Index (SI), 2. cycle period, and 3. number of detected cycles. Permutation tests (α=0.05, p<0.004 with Bonferroni correction) assessed significance, and regression models examined correlations with seizure rate.

Across an average 612-day study period, 22,976 seizures were reported. Following an ASM change, the SI of the seizure cycle was more likely to change (p<0.004). This was pronounced in drug-switching-responders (median absolute SI difference: 0.37 [IQR=0.26] vs. 0.11 [IQR=0.11] in the drug-sustained-responders, p<0.004, permutation test). Changes in cycle length and number of detected cycles were similar across groups, possibly due to a non-linear relationship between seizure rate and cycles, suggested by weak linear correlations and poorly fitting models.

These findings suggest ASMs may influence how strongly seizures synchronise with diary-detected seizure cycles. However, this relationship is complex and not yet well understood, complicating clinical interpretation. Ongoing research into real-time seizure cycle tracking may support the use of seizure cycles in aiding treatment monitoring.

## Introduction

Epilepsy is a chronic neurological disease that causes recurrent seizures and accounts globally for 13 million disability-adjusted life years.^1^ The first-line treatment for epilepsy is antiseizure medication (ASM), which can produce lifelong seizure freedom.^2^ However, more than half of all people with epilepsy undergo years of trial-and-error to find an optimal regime, which in many cases does not achieve seizure freedom.^3^

Assessing ASM efficacy involves titrating to the maximum tolerated dose of a new drug then waiting three times the longest inter-seizure interval or one year, whichever is longest, to determine seizure freedom.^4^ During this ambiguous and lengthy timeframe, patients may make their own efficacy assessments, impacting ASM adherence. Clinicians, in turn, draw on their professional experience to assess therapeutic response earlier, typically by comparing seizure rates before and after treatment.^5^

However, the timing of seizures is not random but exhibits dynamic patterns that may confound frequency-based assessment of treatment efficacy. Most notably, seizures can occur in repeating patterns, known as seizure cycles.^6, 7^ These cycles can emerge across multiple timescales, including daily (circadian), multiday (multidien) and annual (circannual).^8,9^ They are thought to result from various biological rhythms that generate variations in seizure risk, influencing seizure rate.^8, 9^ They have been reported in historical descriptions of epilepsy,^8, 9^ studies of epilepsy colonies,^10–13^ longitudinal human intracranial electroencephalography (EEG) signals,^6, 7, 9, 14, 15^ biosignals recorded by wearable devices,^16,17^ thousands of electronic seizure diaries,^6, 18^ and both naturally occurring canine^19^ and induced rodent models of epilepsy.^20^ Failing to account for seizure cycles when assessing ASM efficacy may lead to inaccuracies, particularly if natural peaks in seizure cycles are mistakenly compared to natural troughs.^5, 21, 22^

Additionally, a prevailing theoretical action of ASM is to reduce seizure rate by raising the seizure threshold. This is based on electroconvulsive therapy studies that required increased electric currents to induce a seizure after ASM administration.^23^ Alternatively, it is plausible that drugs alter cycles of seizure risk relative to the threshold.^24^ More importantly, this could be quantified using the Synchronisation Index (SI),^6^ also known as the Mean Resultant Length or Phase-Locking Value, which indicates how strongly seizures align with a specific cycle (Figure 1).^9^

**Figure 1.**
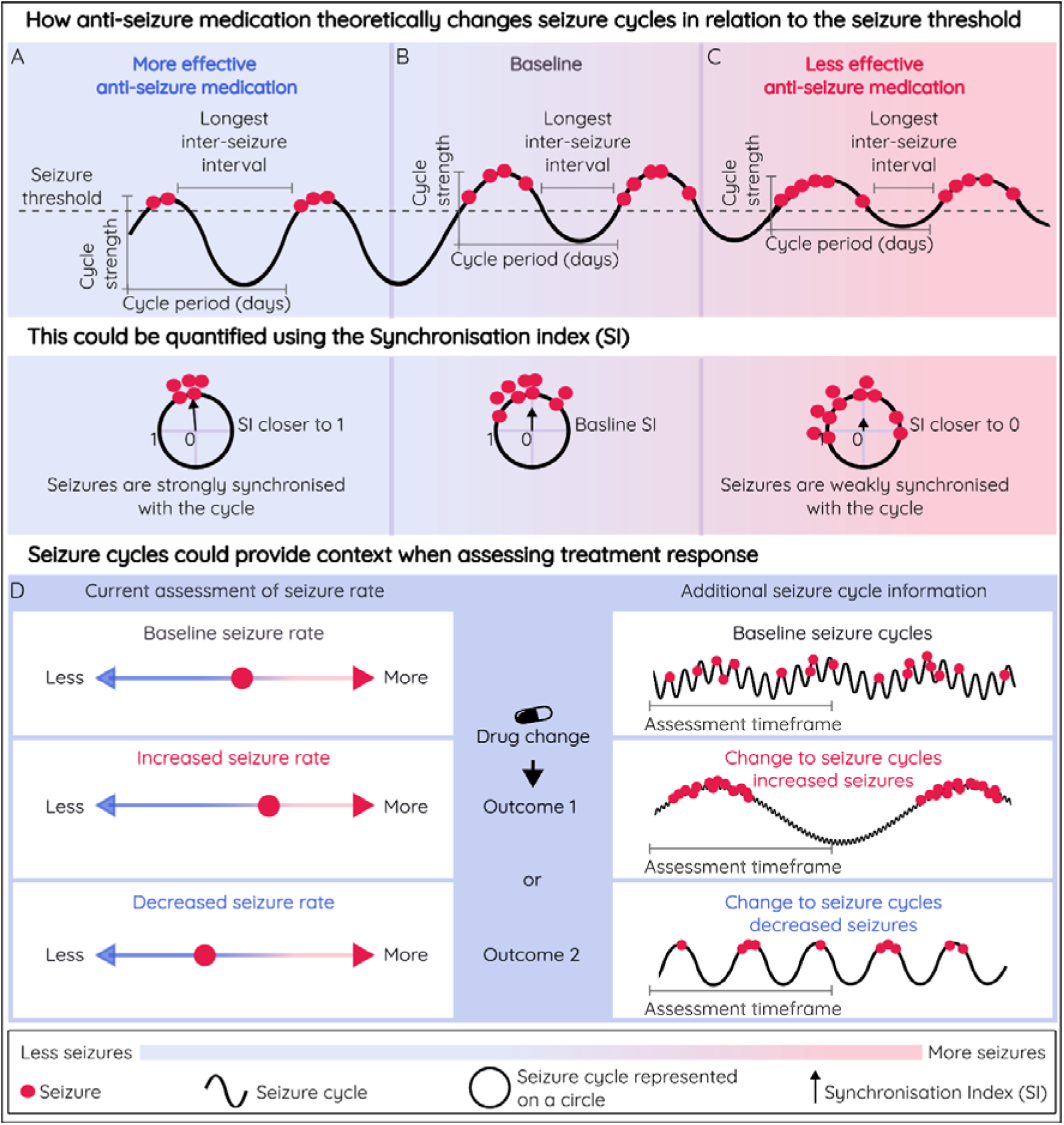
Antiseizure medication may change seizure cycles relative to the seizure threshold, which could be measured by the Synchronisation Index (SI) and used to contextualise changes in seizure rate when assessing treatment efficacy. A seizure cycle could be represented by a sinewave to indicate higher- and lower-risk seizure states that fluctuate over time. The cycle has a specific duration, such as a 10-day period, but varies between a small range of values (e.g. 7-14 days). In this example, the amplitude of the sinewave represents how strongly seizures synchronise with an individual cycle. The strength of the cycle can be estimated by plotting the same seizures on a circle. The average spread of seizures around the circle is represented by the arrow, and the length of the arrow is the SI. The direction of the arrow indicates where in the cycle seizures tend to occur. (A) A seizure cycle predominantly below the seizure threshold compared to baseline (B). (A) The duration of the high-risk seizure phase is reduced, decreasing seizure rate, extending the longest inter-seizure interval and increasing the cycle amplitude or SI. Conversely in (C), a seizure cycle largely above the seizure threshold would increase the duration of time for seizures to randomly occur, increasing seizure rate, reducing both the longest inter-seizure interval and cycle amplitude or SI. Note, at any given time, there could be multiple cycles of seizure risk, with different periods and different relationships to the seizure threshold, all of which would contribute to overall risk and may obscure changes to seizure rate and the longest inter-seizure interval. (D) An example of how monitoring changes in seizure rate and seizure cycles could be used to provide additional information when evaluating treatment efficacy.

The use of seizure timing patterns to enhance ASM evaluation is supported by existing studies investigating the effects of ASMs on seizure cycles,^10–12, 24, 25^ including recent results from a neurostimulation device study^24^ showing effective ASMs reduced both the rates of epileptic activity and their cycle power. However, clinically most treatment decisions rely on assessments of seizure frequency based on self-reported events or seizure diaries.^26^ Therefore, it is equally important to explore the relationship between diary-detected seizure cycles and ASM timing. Fortunately, diary-detected cycles show similar periods to cycles estimated from electrographic seizure times for many patients.^27^

The primary aim of this study was to determine whether ASMs altered patterns of self-reported seizures by measuring differences in seizure cycles before and after an ASM regimen change. If ASMs modify seizure timing, then differences may be measured in the absolute differences in 1. the cycle strength (or synchronisation index; SI), 2. cycle period, and 3. the total number of detected cycles. Analysis of ASMs and seizure cycles using real-world seizure diary data is necessary to, guide further research into how seizure cycle information could optimise ASM timing and support treatment evaluations.

## Methods

### Study Design

This retrospective longitudinal study of two cohorts, reported following STROBE guidelines, compares seizure cycle properties (the strength of the seizure cycle, the duration, and the number of detected cycles) 4 months before and after a drug change. Analyses included whole-group comparisons and sub-group analyses stratified by treatment response at 4 months.

### Participants

Participant with electronic seizure diaries and medication adherence data reported in the commercially available epilepsy management app (Seer app, Seer Medical Holdings Ltd^®^, Australia) were extracted in February 2023.

Inclusion criteria were: (1) use of one or more ASMs during the entire study period; (2) for the drug-switching group, one ASM change after at least 4 months on the first recorded ASM and no changes for 4 months afterward; (3) for the drug-sustained group, no treatment changes during the study period; and (4) a minimum of 4 months of self-reported seizure and medication recordings before and after an ASM change (drug-switching group) or a random timepoint (drug-sustained group). Participants were excluded if they logged the use of an undisclosed medication. There were no inclusion or exclusion criteria based on seizure rate.

### Standard Protocol Approvals and Patient Consents

The Human Research Ethics Committee of St. Vincent’s Hospital, Melbourne approved the use of deidentified mobile application data for this research (LRR 165.19). Mobile application users gave implied consent based on the terms and conditions of use.

### Outcomes

Two clinical outcomes were assessed. First, ASM efficacy was defined as seizure freedom for 1 year or three times the longest inter-seizure interval, whichever was longer, as defined by the International League Against Epilepsy (ILAE).^4^ Second, treatment response was based on a ≥50% reduction in daily seizure rate comparing 4 months before and after a drug change (drug-switching group) or random timepoint (drug-sustained group), aligning with common clinical practice and research standards.^2, 28^

### Missing Data

#### Seizures

Mobile seizure diaries ending before 1 year were treated as missing data. If the longest inter-seizure interval was increasing before the diary ended, the drug was considered effective; if unchanged, ineffective.

#### Antiseizure Medication

If no ASM was recorded, it was unclear whether the participant was not on medication or simply did not log it. Therefore, ASM changes were only confirmed when an initial regimen and a subsequent change (medication or dosage) were recorded in the app.

#### Drug Adherence

Adherence was estimated as the percentage of recorded ‘taken’ doses per day relative to the total doses prompted by the mobile application. To investigate the impact of drug adherence on our analyses, we considered two scenarios: 1) Missing data were treated as completely non-adherent (i.e., all doses were missed), and 2) Missing data were treated as completely adherent (i.e., no doses were missed).

### Seizure Cycle Detection

#### Determining the Best Fitting Cycle Period

A previously described seizure cycle detection and monitoring method^27^ was applied to seizure-onset times from seizure diaries. The method uses the Synchronisation Index (SI), a statistical measure quantifying how closely seizures align with a candidate cycle duration.^6, 7^ SI values range from 0 to 1, where a value of 0 indicates that seizures do not synchronise with the candidate cycle and 1 indicates perfect synchronisation.^12^ Since the number of seizures and seizure rate influence the SI, a minimum of five seizures was required to calculate the SI, and a correction factor was applied.^29^ To identify the most likely seizure cycle period(s), the corrected SI was repeatedly calculated for a range of candidate cycle periods (integers between 1 and 45 days).^7^

#### Selecting Significant Seizure Cycle Periods

To select the most likely cycle period(s) and limit false detections, a permutation test was applied to the corrected SI.^30, 31^ One thousand permutations of the original timeseries were generated by randomly shuffling seizure-onset times with their associated inter-seizure interval.^30^ The corrected SI was computed for each surrogate timeseries and candidate cycle period, to create a period-specific threshold with a 10% false discovery rate.^30–32^ Corrected SI values exceeding the threshold were considered statistically significant and henceforth, referred to as detected seizure cycles.

The maximum number of seizure cycles a person can have is currently unknown. Cyclical seizure models require choosing how many cycle periods to include, with past studies using between 1 and 6 periods.^7, 17, 33, 34^ Setting a universal threshold for selecting influential cycles is challenging due to varying prognostic goals and tolerance for errors. Thus, in this work, the number of significant seizure cycles identified by permutation tests was compared between drug-sustained and drug-switching groups. For further analyses (i.e., comparing changes in corrected SI values and period lengths before and after an ASM change or random timepoint), we considered only the dominant cycle, defined as the statistically significant cycle with the highest corrected SI. Since the dominant cycle period may change (e.g. 4-day cycle to a 25-day cycle), changes in the original dominant cycle (dominant cycle detected before the drug change or random timepoint) were evaluated separately from changes in the new dominant cycle (dominant cycle detected after the drug change or random timepoint).

#### Detecting Seizure Cycles with Variable Durations Using a Sliding Window Approach

To detect cycles that change over time, the corrected SI was calculated for each candidate cycle using a sliding window of data. The window duration aligned with the clinical review period of approximately 4 months and ensured the longest cycle repeated at least three times (e.g., 135 days for a 45-day cycle). The duration was adjusted to be a multiple of the candidate cycle period (e.g., a 132-day window for a 12-day cycle) to prevent bias from incomplete cycles. The window shifted in steps equal to the cycle period to further minimise bias.^27^

#### Seizure Cycle Detection When Medication Becomes Effective

The effect of ASMs on the underlying driver of seizure cycles is unknown. We assume that when ASMs are effective, seizure cycles fall below the seizure threshold and cease to produce seizures. However, it is not yet clear when a presumed inter-seizure interval transitions into a period of seizure freedom. These uncertainties were considered when choosing a sliding window method, which regularly analyses past data to determine current cycle properties. Therefore, to avoid falsely detecting seizure cycles during periods of seizure freedom, cycle results on seizure free days were excluded.

### Statistical Analysis

Demographic and clinical details of drug-switching and drug-sustained groups were compared using the Wilcoxon rank-sum test for continuous non-parametric data, We also used the student’s t-test for continuous parametric data, and the chi-squared test of homogeneity and independence for independent parametric categorical variables.

Permutation tests were performed to evaluate differences in seizure cycle properties, of both the entire cohort and stratified drug-switching and drug-sustained groups. Specifically, permutation tests assessed the absolute difference in seizure cycle properties (SI of both the original and new dominant cycles, dominant cycle period, number of detected cycles) before and after the ASM change or random timepoint.

The permutation test accounts for seizure cycles appearing to be non-stationary, where statistics are approximately constant over an interval of time rather than all time. Normal cycle variation makes it difficult to define a significant or clinically relevant cycle change. Therefore, permutations of the drug-sustained group defined normal cycle variability (null hypothesis) as any seizure cycle change occurring after a random timepoint. The random timepoint occurred after the first 4 months of data and was created using a uniform pseudorandom number generator with a fixed seed. The process was repeated 1000 times per drug-sustained participant. The absolute difference in cycle properties (SI of the original and new dominant cycles, dominant cycle period, number of detected cycles) and daily seizure rate before and after the random or ASM timepoint was calculated for each group. Median values were derived from each permutation of the drug-sustained group. This established a distribution of medians under the null hypothesis. This null distribution was used to compare the drug-switching group’s median and compute a p-value. Bonferroni correction for multiple comparisons with α=0.05 was applied, giving a significance threshold of p=0.004.

To further investigate the relationship between seizure rate, which defines treatment response, and seizure cycle properties, values from both cohorts were visually inspected using scatter plots. Linear regression was applied to estimate their correlation. The adjusted-R^2^ value was used to measure model fits. All analyses were undertaken using MATLAB v2024b and the Circular Statistics Toolbox.^35^

## Results

### Participants

A total of 86 participants were included after exclusions, with 45 (52%) undergoing ASM changes and 41 (48%) on stable antiseizure medications (ASM). The mean age was 34.89 years (SD = 17.11). There were 49 females, 35 males and 1 non-binary person. Sixteen people had generalised epilepsy and 46 had focal epilepsy (Supplementary Table 1). Across all participants, 22,976 seizures were recorded. The drug-switching and drug-sustained groups had comparable demographics, seizure rates, diary durations (median ∼254 vs. 214 days), and ASM adherence, with no significant differences in these covariates. No participants achieved complete seizure freedom. The number of responders (n = 7/45 vs. n = 8/41) and use of mono-versus polytherapy also did not differ significantly between groups. Drug-switching group participants had a higher likelihood of using comorbid medications (p < 0.004). Detailed demographic data and additional group characteristics are provided in the supplementary material (supplementary Tables 2-6 and supplementary Figure 1).

### Antiseizure Medications and Seizure Timing

#### Synchronisation Index (SI)

One or more seizure cycles were identified in all participants, but not at every timepoint. No significant change was found in the SI of the original dominant cycle (median absolute SI difference: 0.07 [IQR = 0.27] in the drug-switching group vs. 0.06 [IQR = 0.02] in the drug-sustained group; p = 0.25, permutation test; Figure 2A).

**Figure 2.**
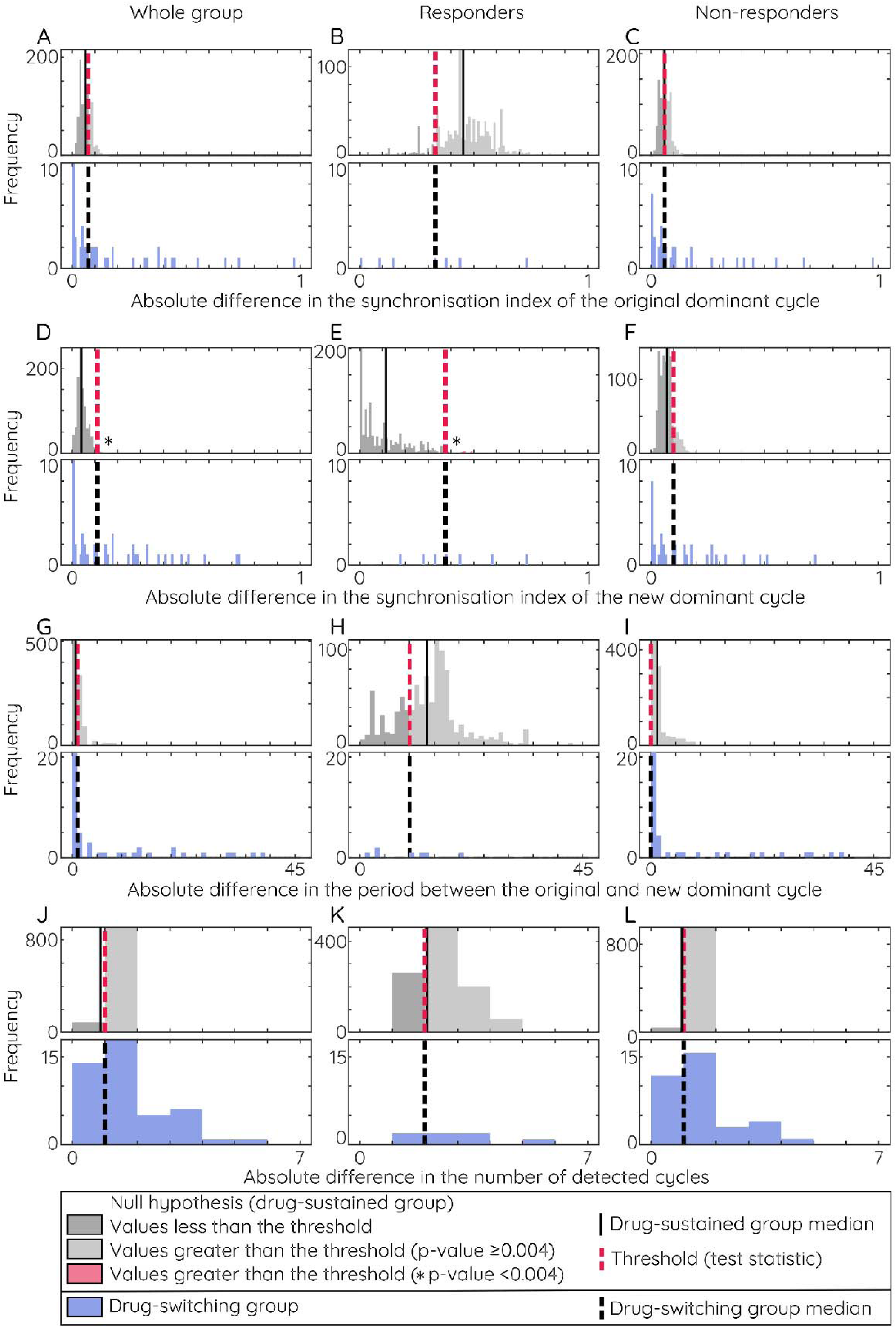
Permutation test result: absolute difference in seizure cycle variables 4-months after a change in antiseizure medication (ASM) or random timepoint. To assess whether seizure cycles changed after an antiseizure medication (ASM) adjustment, participants who changed ASMs (drug-switching group) were compared to those who remained on the same medications (drug-sustained group) using a permutation test. A null distribution (grey histograms) was generated from the drug-sustained group by randomly sampling timepoints from each participant’s seizure diary and calculating the absolute difference in seizure cycle variables before and after, then computing the median absolute difference. Variables included the Synchronisation Index (SI) of the original and new dominant cycles, cycle period, and number of detected cycles. The average difference in the drug-switching group (black dashed line) served as the test statistic (pink dashed line), with the p-value calculated as the proportion of the null distribution exceeding this threshold—shown as a light grey histogram (not significant) or pink histogram (significant) to the right of the pink dashed line. Figure 2D shows a significant change in the SI of the new dominant cycle following ASM change. This effect was also seen in participants with ≥50% seizure reduction at 4 months (Figure 2E). No other variables showed significant changes.

In contrast, a significant change in the SI of the new dominant seizure cycle was observed when comparing the drug-switching group to the drug-sustained group (median absolute SI difference: 0.11 [IQR = 0.28], n = 45 vs 0.04 [IQR = 0.02], n = 41, respectively, p < 0.004, permutation test; Figure 2D). This difference occurred despite all participants being on ineffective medication, according to the ILAE definition.

When stratified by treatment response at 4-months, a ≥50% reduction in seizure rate was associated with a greater change in the SI (Figures 2B and 2E) – particularly for the new dominant cycle (median absolute SI difference: 0.37 [IQR=0.26] in the drug-switching-responders (n = 7/45) vs. 0.11 [IQR = 0.11] in the drug-sustained-responders (n = 8/41), p < 0.004, permutation test).

Among non-responders (<50% seizure rate reduction at 4-months), only small absolute SI differences were observed. While the drug-switching group showed greater variability, differences between groups were not significant (Supplementary Table 7). For the original dominant seizure cycle, the median absolute SI difference was 0.06 [IQR=0.16] in drug-switching-non-responders (n = 38/45) vs. 0.06 [IQR=0.02] in drug-sustained-non-responders (n = 33/41; p = 0.46, permutation test; Figure 2C). For the new dominant cycle, the difference was 0.10 [IQR=0.25] vs 0.07 [IQR=0.03], respectively (p = 0.13, permutation test; Figure 2F).

#### Dominant Cycle Period

On average, the dominant cycle period did not change (Figure 2G, median values near zero). However, changes in the period were more likely when ASMs changed (Figure 2G, lower panel – note the left-skewed distribution) and also, when there was ≥50% reduction in seizure rate regardless of an ASM change (Figure 2H, median values shifted to the right with a broader distribution). However, the changes in cycle period were not statistically significant (Supplementary Table 7).

#### Number of Detected Seizure Cycles

In both drug-switching and drug-sustained groups, there were up to six detected seizure cycles (Figure 2J-L). The number of detected cycles changed by 1–2 cycles on average (see Figure 3 for case examples). No significant differences were observed between groups (Supplementary Table 7).

**Figure 3.**
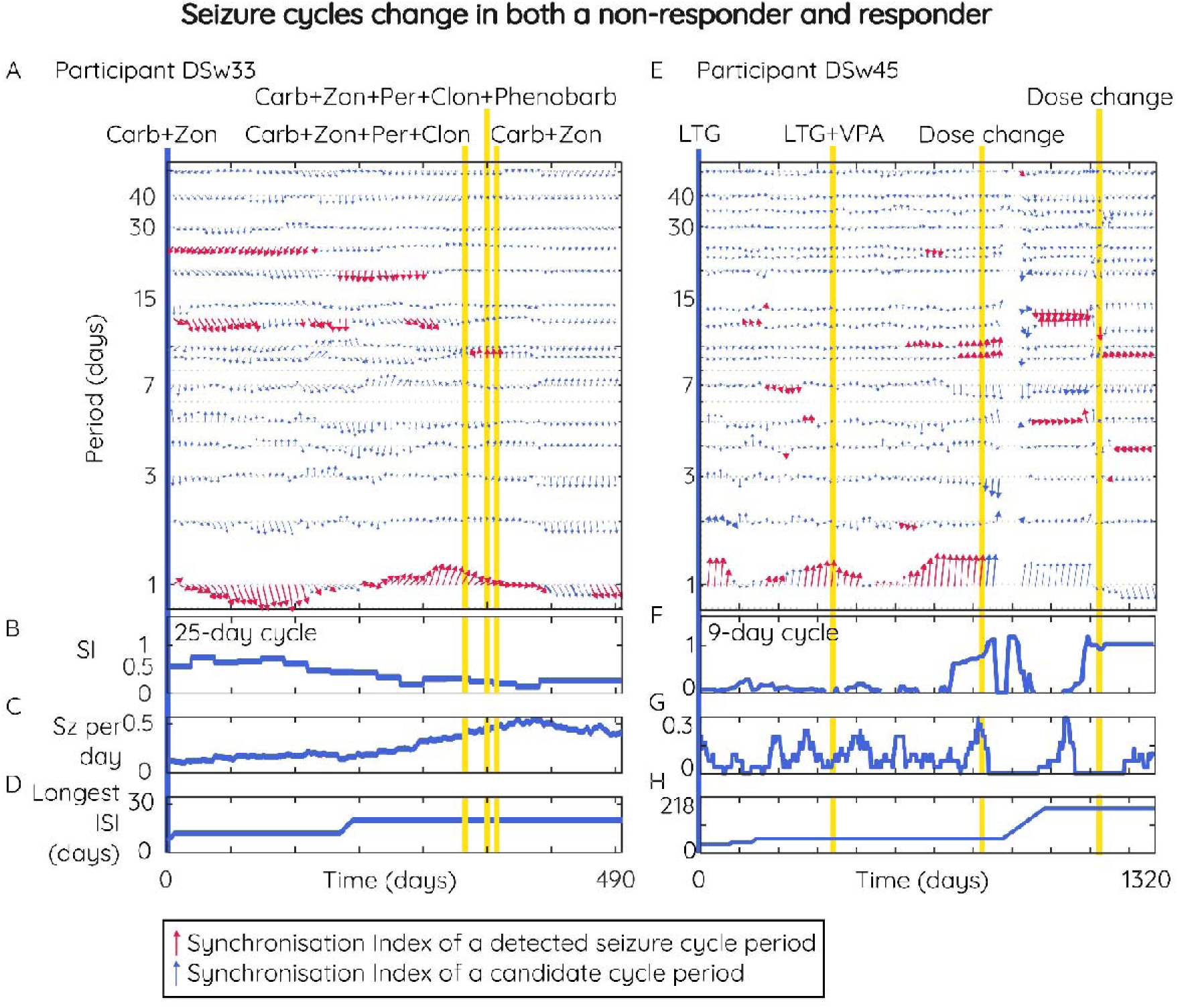
Case examples of seizure cycles changing in a non-responder and responder from the drug-switching group. (A) Participant DSw33 (non-responder): Vector plot of detected (i.e., significant) seizure cycles (red arrows). Multidien seizure cycles were present for 322 days from study onset (x-axis) but disappeared after changing antiseizure medications (ASM; yellow vertical lines). The circadian cycle is detected for most of the 490 days. (B) The SI of the 25-day seizure cycle decreases over 290 days. (C) Seizure rate continues to increase after the change in antiseizure medication. (D) The longest inter-seizure interval does not triple. (E) Participant DSw45 (responder): Vector plot of detected seizure cycles. A circadian seizure cycle was detected for 900 days from study onset. During this time, about-weekly and about-fortnightly cycles were intermittently detected. After the first medication dose change, the circadian cycle was no longer detected, but the about-weekly and about-fortnightly cycles persisted. (F) The SI of the 9-day seizure cycle decreased then increased around drug changes. (G) After a dose adjustment in ASM, the seizure rate decreased, accompanied by the emergence of a possible multi-month seizure cycle, which went undetected due to the duration of the data and method specifications. (H) The longest inter-seizure interval tripled but remained under a year. Note, for vector visualisation, the y-axis of the vector plot is on a logarithmic scale. Along the x-axis, vectors (representing the SI computed over the previous 4 months) are down sampled and plotted every 2 weeks in Figure 2A and every 3 weeks in Figure 2E. Carb: carbamazepine, Clon: clonazepam LTG: lamotrigine, Per: perampanel, SI: Synchronisation Index, Sz: seizures, VPA: valproate, Zon: zonisamide, DSw: drug-switching group.

#### The Relationship Between Seizure Rate and Seizure Cycles

The fact that similar changes in cycle variables were seen in both responders and non-responders in both the drug-switching and drug-sustained groups suggests that these cycle features may reflect general patterns related to seizure rate, rather than treatment response alone. However, regression analysis indicates that no linear correlation exists between seizure rate and the three cycle properties (Figure 4).

**Figure 4.**
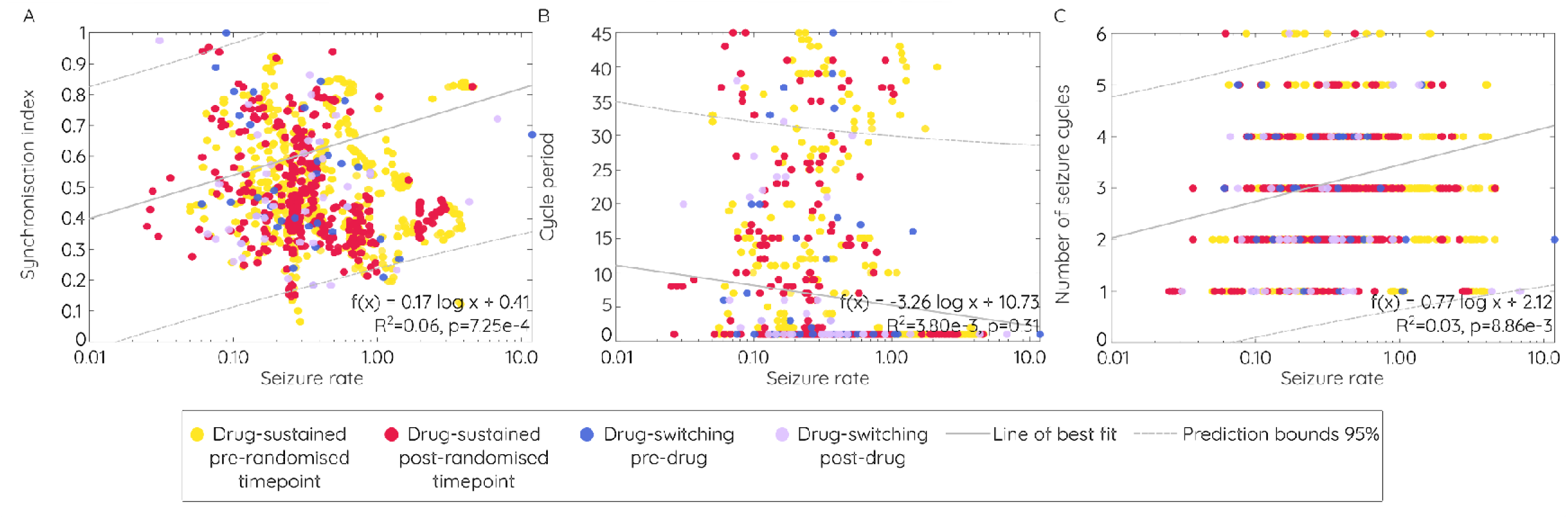
No linear correlation exists between seizure rate and seizure cycle variables. Seizure rate is plotted on the x-axis (log scaled) against cycle variables in (A) the synchronisation index of the original dominant cycle period (B) the dominant cycle period and (C) the number of detected seizure cycles. For each variable, colours indicate groups and before or after the anti-seizure medication (ASM) change or random timepoint: before the ASM change (blue), after the ASM change (purple) for the drug-switching group: before a randomised timepoint (yellow) or after a randomised timepoint (red) for the drug-sustained group.

Instead, seizure rates that are similar for clinical interpretation – though not identical in value (e.g. 44.5 versus 45.6 seizures per 4 months) – appear to exhibit a non-linear, one-to-many relationship with seizure cycle properties, independent of treatment or response. This relationship is evident visually, as a range of dominant cycle periods (1–45 days) with SI values between 0.18 and 1.00 and number of detected cycles (1–6) are associated with comparable seizure rates (see Supplementary Figure 2 for details).

## Discussion

### Overview

This study assessed changes to seizure timing following ASM adjustments using an electronic seizure diary-based approach, thereby providing a novel and practical benchmark to guide cycle-based clinical assessment. All participants were on ineffective ASMs at some point during the study, with only a few – across both drug-switching and drug-sustained groups - classified as responders at 4 months. The results provide preliminary evidence that ASMs may alter self-reported seizure cycles beyond natural variability. Our findings suggest that antiseizure medications ASMs can influence the underlying temporal organization of seizures, specifically by altering how strongly seizures synchronize to a cycle (Synchronisation Index, SI), particularly if the ASM produces a reduction in seizure rate. This observation extends beyond natural variability, suggesting a possible pharmacodynamic effect on seizure timing. While ASMs did not significantly impact the dominant cycle period or the number of detected cycles, the change in SI presents a novel, albeit complex, metric for understanding treatment response that should be investigated further in future work.

### The Relationship Between Antiseizure Medication, Seizure Rate and Cycles

Medication changes were more likely to affect how strongly seizures synchronise with a cycle (as measured by the absolute change in the SI). However, the currently unpredictable relationship between seizure cycles and seizure rate means changes in cycle synchronisation warrant further investigation as an indicator of drug efficacy for clinical decision-making. Of note, self-reported seizure rates are an unreliable biomarker,^36, 37^ whereas self-reported seizure cycles may align more faithfully to electrographic seizures.^27^ This finding is supported by inconsistent evidence related to the relationship between seizure rate and the SI. For instance, a study of a small cohort of drug-resistant focal epilepsy with an intracranially implanted Neuroresponsive Stimulation (RNS) system (NeuroPace, Mountain View, CA) found a negative linear correlation between seizure rate and the SI (not corrected) from multidien cycles, but no such relationship for circadian seizure cycles.^7^ In contrast, a study of a large electronic seizure diary cohort reported a negative correlation between seizure rate and both circadian and multidien cycles, but did not specify the exact nature of the relationship.^6^

The current findings share similar SI and seizure rate distributions with the two referenced studies,^6, 7^ and suggest the relationship between seizure rate and the SI may be non-linear and one-to-many. Mathematically, a seizure rate of 0.10 could arise from approximately 1 seizure every 10 days (about-weekly cycle), 3 seizures every 30 days (about-monthly cycle), or 36 seizures over 365 days (a circannual cycle). Given the infinite possible combinations, it seems inevitable that individuals with different underlying cycle patterns could exhibit the same average seizure rate. This creates a complex relationship between seizure rate and seizure timing, warranting further investigation using alternative non-linear methods and objectively monitored seizure cycles.

The only other modern study to investigate ASMs and cycles in epilepsy used the RNS system, which records cycles of interictal epileptiform activity (IEA) that synchronise with seizure cycles.^24^ They found that ASMs that reduce seizure rates had no effect on the period of the IEA cycle or the strength of seizures synchronising to the IEA cycle. However, there was a decrease in the strength of the IEA cycle as measured by wavelet power.^24^ Notably, a drug related decrease in seizure rate was not always accompanied by a decrease in IEA cycle strength.^24^ This finding may align with the possibility of multiple seizure cycle generators. That is, if multiple biological rhythms – one of which is represented by IEA cycles – exerts influence over seizure risk, then a reduction in seizure rate without a reduction in IEA strength might suggest the ASM influenced a different biological rhythm.

However, prior RNS research shows ASMs reduce IEA rates,^38, 39^ which impacts cycle detection methods like wavelet power. Controlling for IEA rates may uncover different effects of ASM on IEA and seizure cycles than previously reported. Additionally, RNS stimulation settings can lower both wavelet power of IEA cycles and the SI.^40, 41^ While Friedrichs-Maeder et al. (2024) controlled for individual stimulation changes, group-level differences may have confounded results.

Nevertheless, it is possible that ASMs influence seizure timing through other mechanisms besides rates of seizures or IEA. Hopkins et al. (1985) created a periodic seizure model and demonstrated possible effects of ASMs on periodicity by altering seizure rate and/or the likelihood of transitioning between low to high or high to low levels of seizure risk.^5^ Thereby exhibiting three ways ASMs influence cycles.^5^

It is also possible that side effects of different ASMs may have different effects on seizure timing. Valproate has been studied the most in relation to the circadian cycle, and has been shown to alter circadian CLOCK-gene expression *in vitro*^42, 43^ and in developing rodents exposed to valproate *in utero.*^44^ Valproate also lengthens the period and can cause aperiodicity of the circadian sleep-wake cycle in drosophila.^45^ It also affects the circadian seizure cycle through circadian fluctuations in pharmacokinetics observed in rodents.^46^ This is thought to be due to circadian variations in hepatic blood flow, metabolic enzymes, and plasma protein binding.^46^ Although, it is unclear how these factors would extrapolate to multidien cycles. Possible influences on monthly seizure risk like neurosteroids, also affect valproate serum levels,^47^ but are insufficiently explored regarding their interaction with seizure timing.^48^ However, it is interesting to consider that different drugs with various mechanisms of action, could have different effects on the rhythms of the brain. Future research could reanalyse longitudinal seizure diary data from randomised controlled drug trials to further disentangle this topic.

### Limitations of Using Paper-Based Methods to Monitor Seizure Cycles for Timing Medication and Drug Efficacy Assessments

Seizure cycles are conceptually simple but complicated by the presence of multiple, patient-specific cycles that vary in duration and may change with medication. This poorly understood variability will likely make it difficult to infer seizure cycles using seizure diaries—especially when relying on paper-based methods rather than advanced time-series analysis—to guide medication timing (e.g., chronotherapy, the scheduling of ASMs during periods of increased seizure risk) or to evaluate treatment. This issue is exemplified in the only two studies that attempted chronotherapy^48, 49^ and menstrual phase aligned ASM assessments^48^ for people with presumed catamenial epilepsy. These studies^48, 49^ assumed the phase of the seizure cycle was determined by the onset of menses. However, based on large epidemiological studies, it is well known that normal regular menstrual periods vary within individuals by 7–9 days and the amount of variability is age specific.^50^ Unfortunately, intrasubject period variability was not considered by Feely et al., (1982) and several women received ASM later than planned.^49^ Moreover, although the randomised controlled trial of adjunctive cyclical progesterone therapy by Herzog et al. (2012) accounted for menstrual period variability,^48^ it may not have aligned with seizure cycle variability.^51^ This mismatch may explain why only a sub-cohort benefited from chronotherapy.^48^ Thus, quantifying natural seizure cycle variability through extensive epidemiological studies, could help to improve paper-based cycle monitoring, which would have less barriers than a cycle-tracking algorithm to translate into clinical practice. However, a seizure cycle tracker, akin to a cycle-based seizure forecaster,^52–56^ would probably be more reliable to support clinicians and patients.

### Limitations

This study encountered multiple constraints including missing demographic data, the influence of mobile phone application usage, sampling bias, small sample size (mitigated using permutation tests), self-reporting biases, and inaccurate reporting in electronic seizure diaries (although, more accurate than paper-based diaries^57^). The issue of inaccurate self-reported seizures is well known, yet diaries are still used to assess drug efficacy in both clinical and research settings^58^ and research to investigate its influence on seizure cycle detection is ongoing.^27^ Despite the limitations of diaries, our results are consistent with previous studies on seizure cycles using intracranial EEG.^24^ Furthermore, although diary-based cycle detection may only be possible in individuals who can precisely record seizures, its value lies in the future possibility of providing extra information to a broader group of patients, to improve treatment decisions.

Since diaries are more likely to have reduced accuracy for nocturnal seizures,^37^ this limitation hindered investigations into clinically relevant changes of the circadian cycle phase. For example, shifts from nocturnal seizures to diurnal seizures may be significant for certain types of epilepsy, such as juvenile myoclonic epilepsy, childhood epilepsy with centrotemporal spikes, and frontal lobe epilepsy.^59^ However, previous studies of epilepsy colonies found some ASMs (of that time) suppressed seizures shortly after administration.^11^ However, they also found other ASMs caused a transition in 24-hr seizure patterns from clustered to dispersed,^11^ and two different ASMs had no association between administration times and daily seizure rhythms.^10, 12^ To progress our understanding of ASM effects on circadian seizure cycles, cycle detection methods could use ultra-long-term sub-scalp EEG^34, 60^ or wearables devices^17, 61, 62^ to record seizures.

Drug non-adherence affecting therapeutic ASM levels may have also obscured results. The absence of a definitive method to measure drug adherence^63^ complicates this assessment. However, both drug-sustained and drug-switching groups reported similar adherence, which was higher than other studies due to drug adherence mobile notifications.^64^ Moreover, in rats, sub-therapeutic levels of levetiracetam due to non-adherence did not affect diurnal seizure patterns.^65^

It is also possible that infections and sleep deprivation—indicated by the use of antimicrobials and hypnotics, respectively, in the drug-switching group—may have influenced seizure timing. These known seizure triggers represent potential confounders that could not be addressed in this study but would be important to investigate in future work. Although the drug-sustained group and permutation test mitigated biases from most variables, group differences in comorbid medications directs future research towards elucidating the effects of comorbidities and drug-drug interactions on seizure timing.

### Conclusion

In conclusion, the results from this study suggest that ASMs can indeed influence the synchronisation strength of seizure cycles, suggesting a direct effect on the temporal organisation of epilepsy. In particular, the observed one-to-many relationship, where similar seizure frequencies can manifest from diverse underlying cycle patterns, challenges the conventional reliance on seizure frequency as the primary indicator of treatment success. Thus, integrating seizure cycle information may provide a more nuanced interpretation of reported changes in seizure frequency after treatment changes, thereby facilitating more precise and personalised therapeutic interventions for people with epilepsy.

## Data availability

Deidentified electronic seizure diary data are available upon reasonable request to co-author Prof. Mark J. Cook.

## Acknowledgments

We thank Michaela Vranic-Peters for her advice regarding the statistical methods used in this study.

## Author contributions

A.R. undertook the literature review, study design, data analysis, results interpretation, created the figures and tables, contributed to concepts, wrote and edited the manuscript. R.E.S. assisted with the study design, results interpretation and edited the manuscript. S.H. designed participant sampling, collected and undertook preliminary data analysis, editing the manuscript. P.K. obtained ethics approval, assisted with results interpretation, contributed to concepts and edited the manuscript. E.N. provided access to the data, contributed to concepts, edited the manuscript and supervised S.H. Authors A.P., A.L., and D.B.G assisted with the study design, results interpretation, contributed to concepts, edited the manuscript and supervised A.R. Author M.J.C. contributed to concepts, created and provided access to the data, results interpretation, designed figure 1D, edited the manuscript and supervised A.R. All authors approved the final version of the manuscript. A.R., S.H., R.E.S., P.K., E.N., and M.J.C. had full access to the data. No author was precluded from accessing the study data.

## Supplementary materials

### Participants

There were 10,015 application downloads as of February 2023; 327 people had drug adherence data, and of those, 87 had ≥4 months of seizure diary data. One participant was excluded because they were taking a non-disclosed research trial medication, leaving a total of 86 participants. We infer the possibility that participants might be living outside of the study location (Melbourne, Australia) and come from any socio-economic background given the application was globally available on >95% of mobile devices. However, the app and therefore the participants, were limited to English-speaking people.

**Supplementary Table 1.**
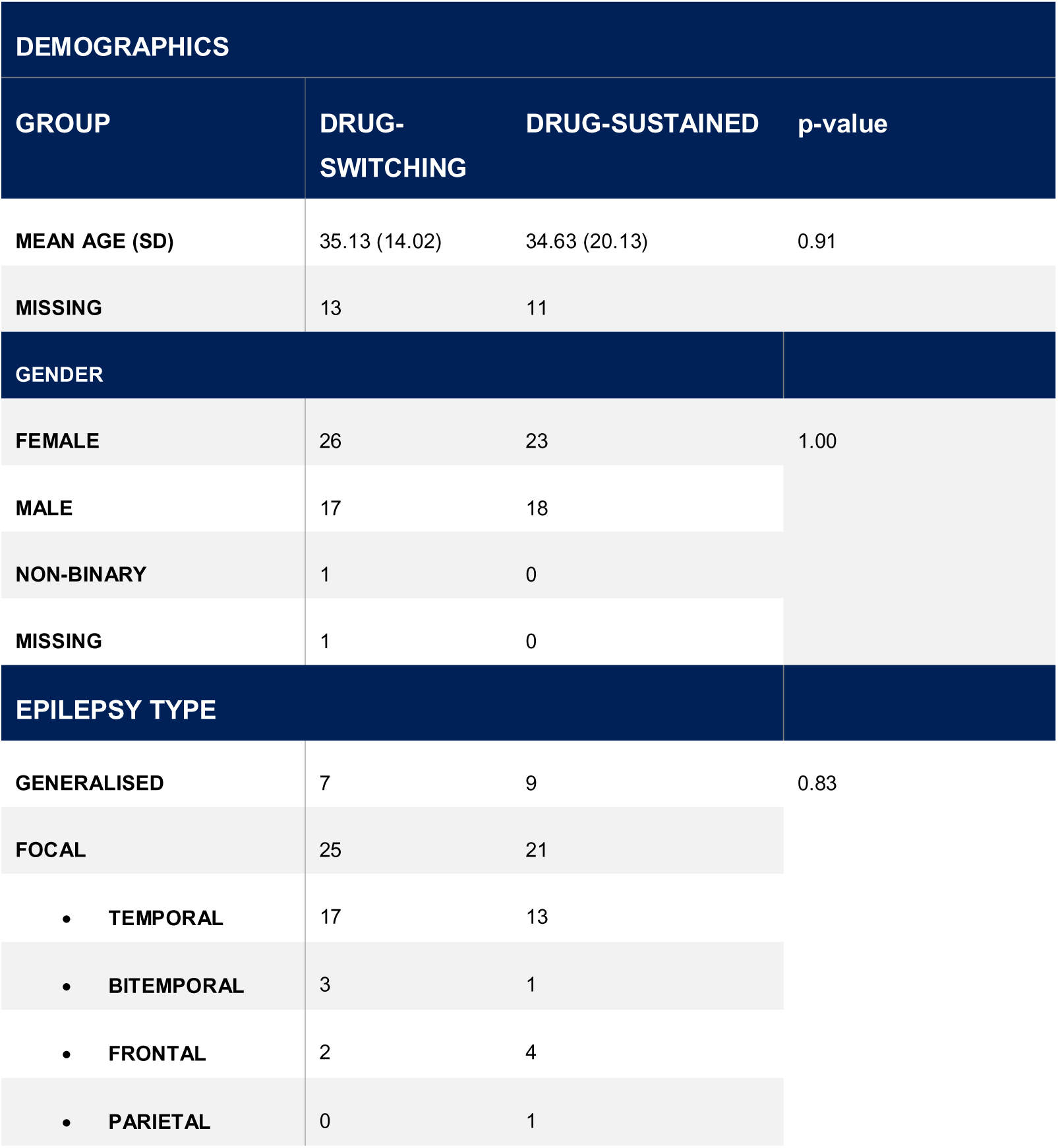
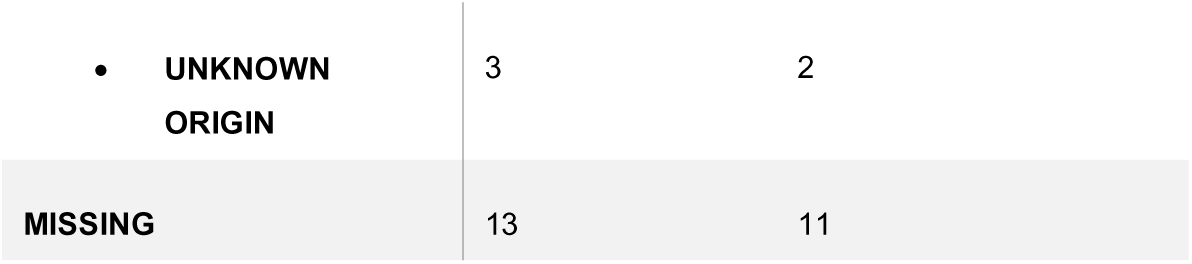
Demographic information.

| DEMOGRAPHICS |  |  |  |
| --- | --- | --- | --- |
| GROUP | DRUG-SWITCHING | DRUG-SUSTAINED | p-value |
| MEAN AGE (SD) | 35.13 (14.02) | 34.63 (20.13) | 0.91 |
| MISSING | 13 | 11 |  |
| GENDER |  |  |  |
| FEMALE | 26 | 23 | 1.00 |
| MALE | 17 | 18 |  |
| NON-BINARY | 1 | 0 |  |
| MISSING | 1 | 0 |  |
| EPILEPSY TYPE |  |  |  |
| GENERALISED | 7 | 9 | 0.83 |
| FOCAL | 25 | 21 |  |
| • TEMPORAL | 17 | 13 |  |
| • BITEMPORAL | 3 | 1 |  |
| • FRONTAL | 2 | 4 |  |
| • PARIETAL | 0 | 1 |  |

|  |  |  |
| --- | --- | --- |
| • UNKNOWN ORIGIN | 3 | 2 |
| MISSING | 13 | 11 |

### Antiseizure Medication, Seizure Rate and Covariates

Forty-one (48%) participants were on stable ASMs and 45 (52%) participants changed ASMs (Supplementary Table 2 and 3), although none were effective in producing complete seizure freedom. In total, there were 22,976 seizures reported (drug-sustained group: 8,313, Median = 92, IQR = 157, drug-switching group: 14,663, Median = 129, IQR = 184.75). There were no significant differences between drug-sustained and drug-switching group daily seizure rates after one drug-switching group outlier was removed due to a high daily seizure rate that was unmatched in the drug-sustained group (drug-sustained group: median daily seizure rate 0.22, IQR = 0.28, drug-switching group: median daily seizure rate 0.24, IQR = 0.31, p = 0.71, Wilcoxon rank-sum test). There was no significant difference in the change in daily seizure rate (drug-sustained group: median difference in seizure rate before and after a random timepoint was 0.06 seizures per day, 95% CI [0.03, 0.10], drug-switching group: median difference in seizure rate before and after treatment was 0.01 seizures per day, p=0.99, permutation test; Supplementary Figure 1) after the one outlier was removed. The number of responders was similar between the drug-switching (n=7/45) and drug-sustained groups (n=8/41).

**Supplementary Figure 1.**
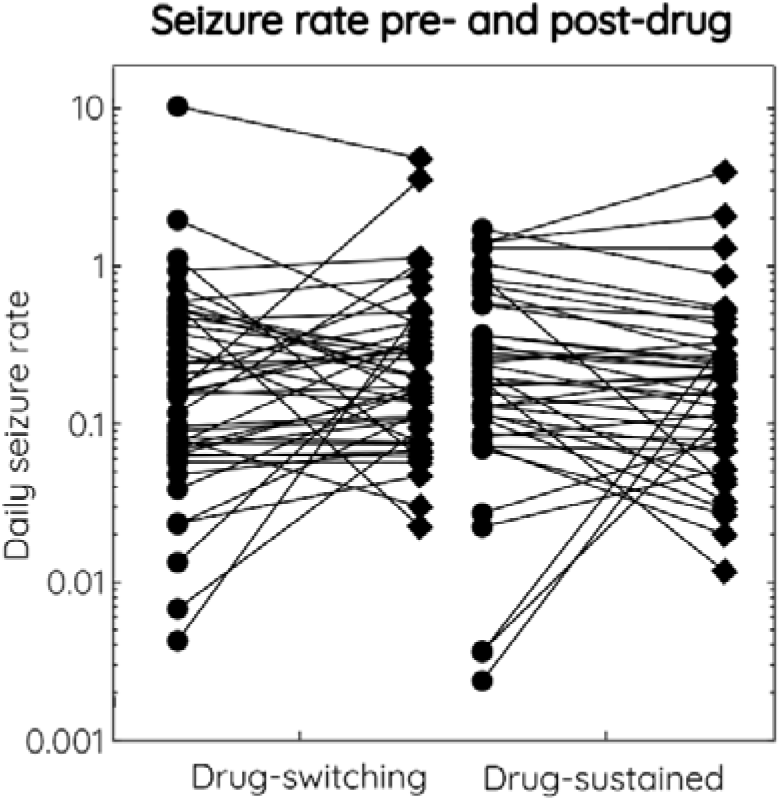
The change in seizure rate before and after an anti-seizure medication change (drug-switching group) or random timepoint (drug-sustained group). Circles symbolise the seizure rate before, and diamonds symbolise after. Note, both x- and y-axes are logarithmically scaled.

There were no significant differences in diary duration (drug-sustained group: Median = 214 days, IQR = 115.75 days, drug-switching group: Median = 254 days, IQR = 323 days, p = 0.65, Wilcoxon rank-sum test), number of participants using mono-or polytherapy (Supplementary Table 4), ASM class (Supplementary Table 5), or ASM adherence (drug-sustained group: when missing data was treated as completely non-adherent Mean = 0.34, SD = 0.42, treated as completely adherent Mean = 0.54, SD = 0.43; drug-switching group: completely non-adherent Mean = 0.47, SD = 0.40, or completely adherent Mean = 0.55, SD = 0.44, p = 0.29, student’s t-test) between drug-sustained and drug-switching groups. However, drug-switching group participants were more likely to use comorbid medications, which were significantly different (p<0.006, chi-squared test) compared to the drug-sustained group (Supplementary Table 6).

**Supplementary Table 2.**
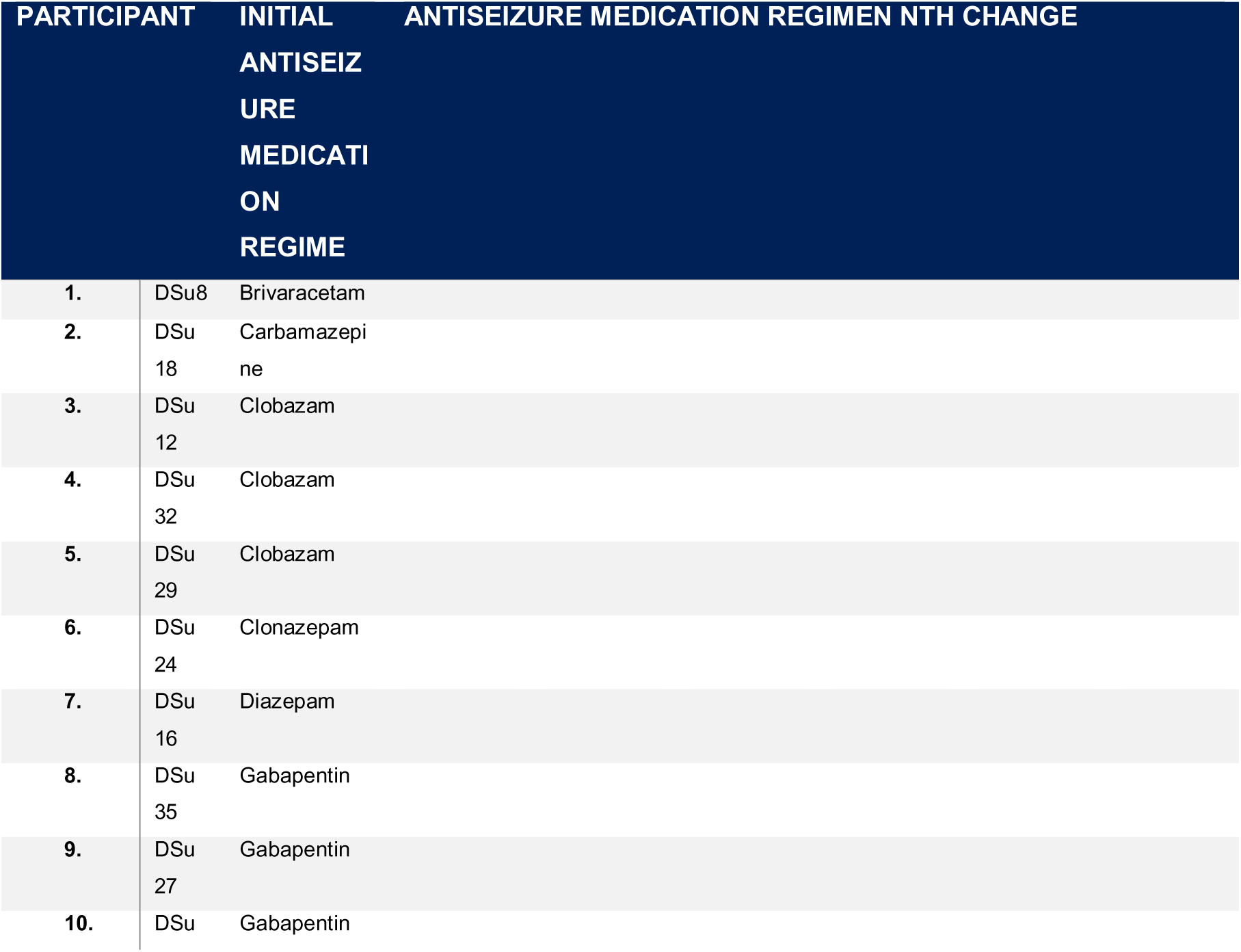
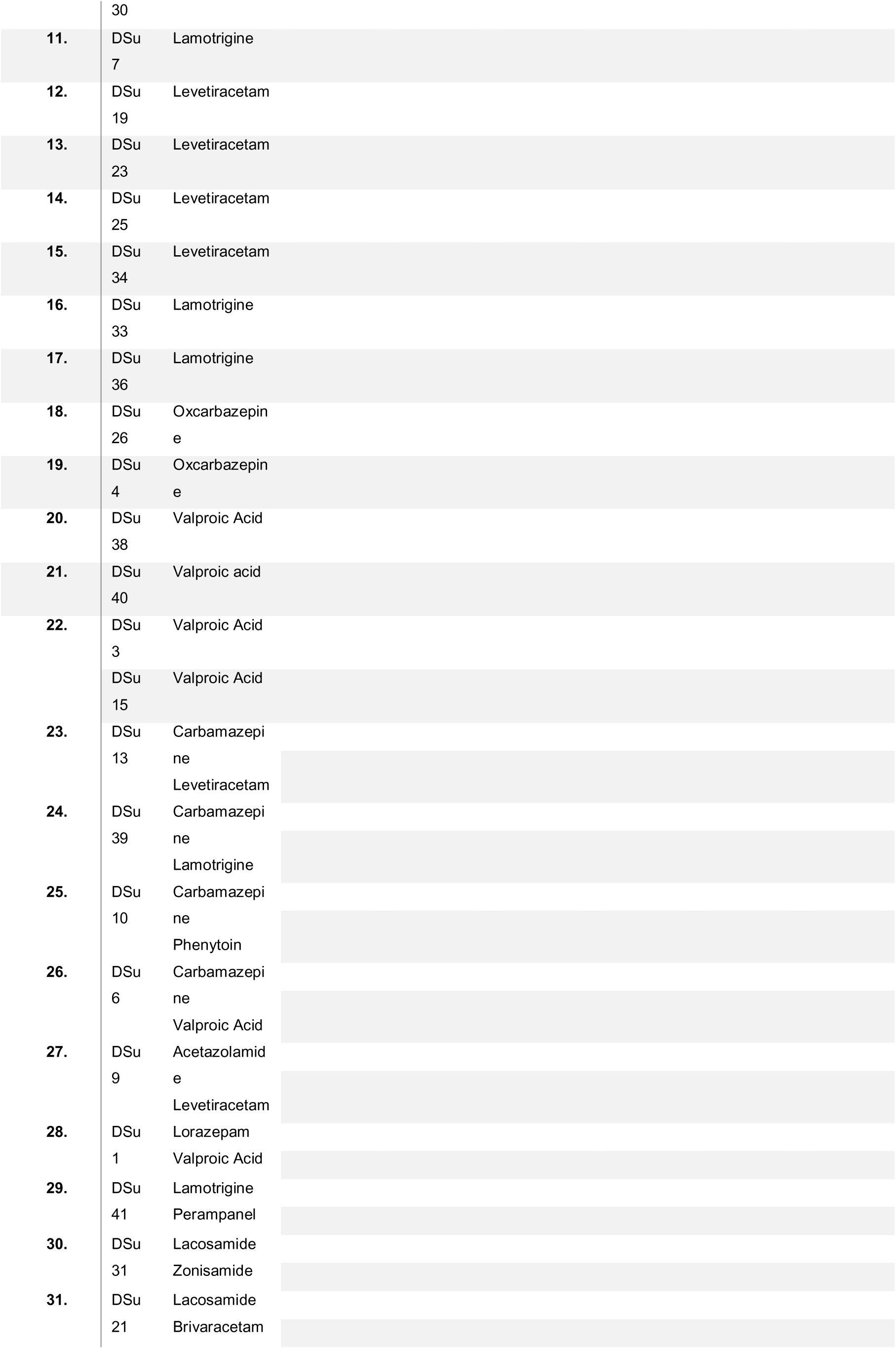
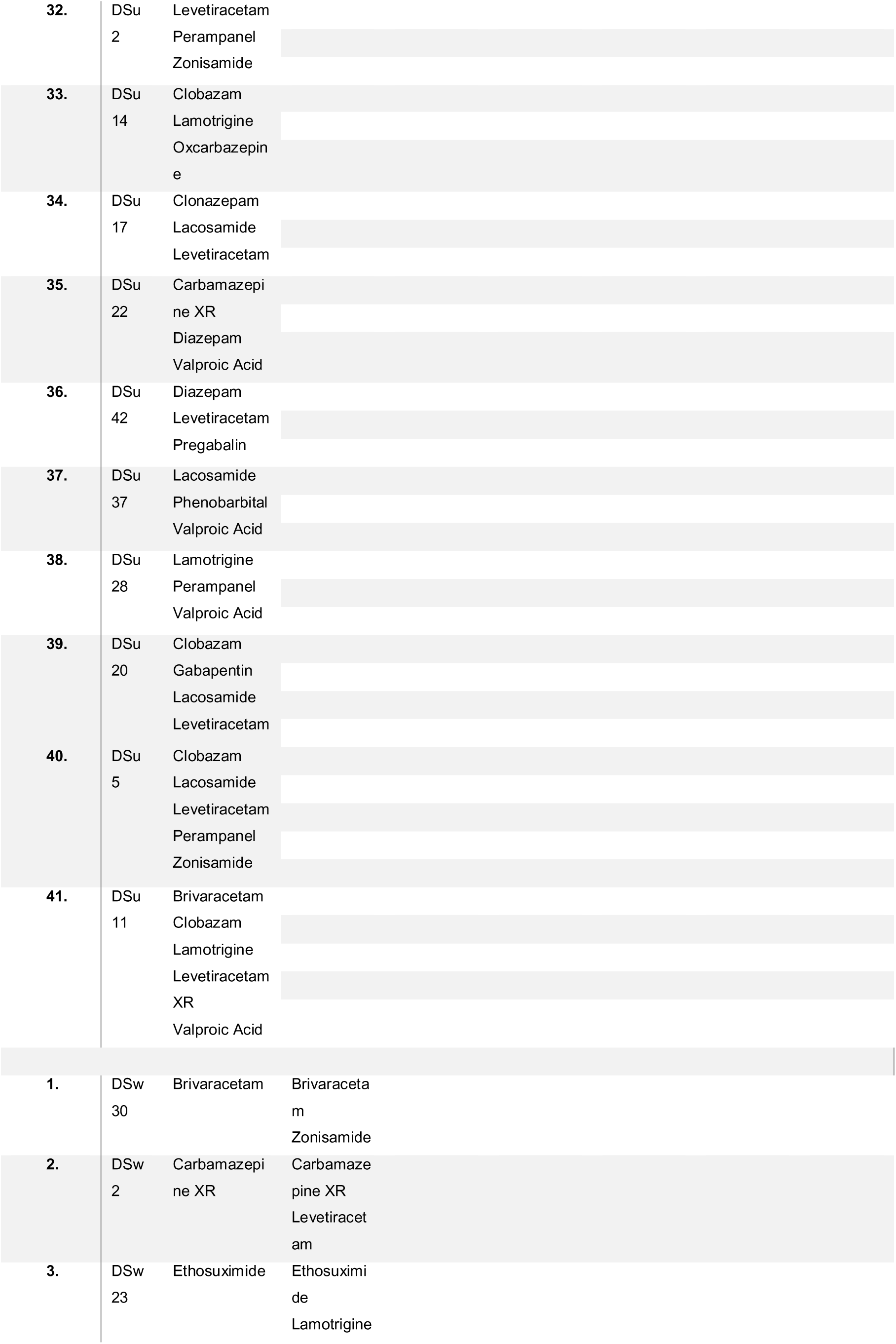
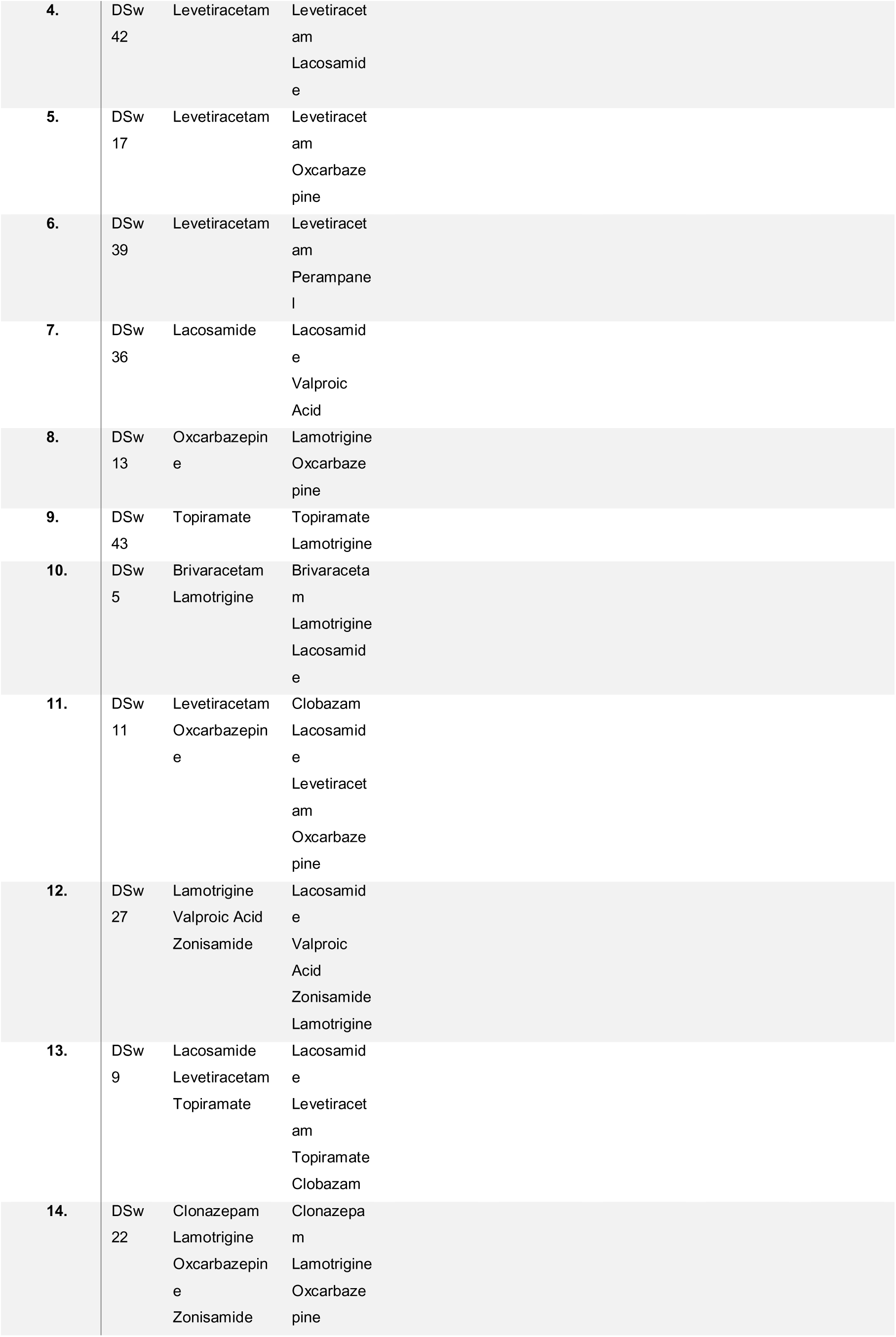
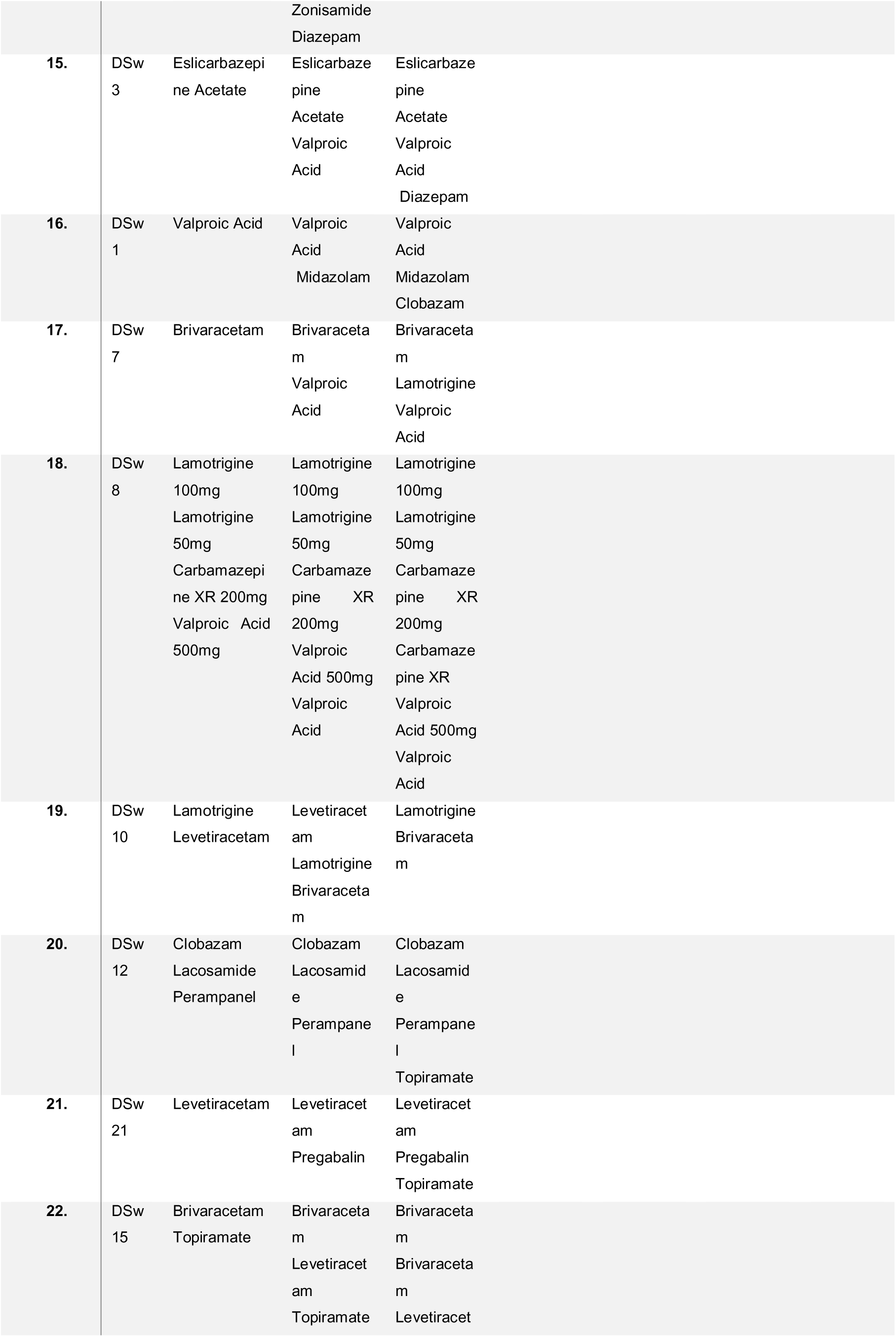
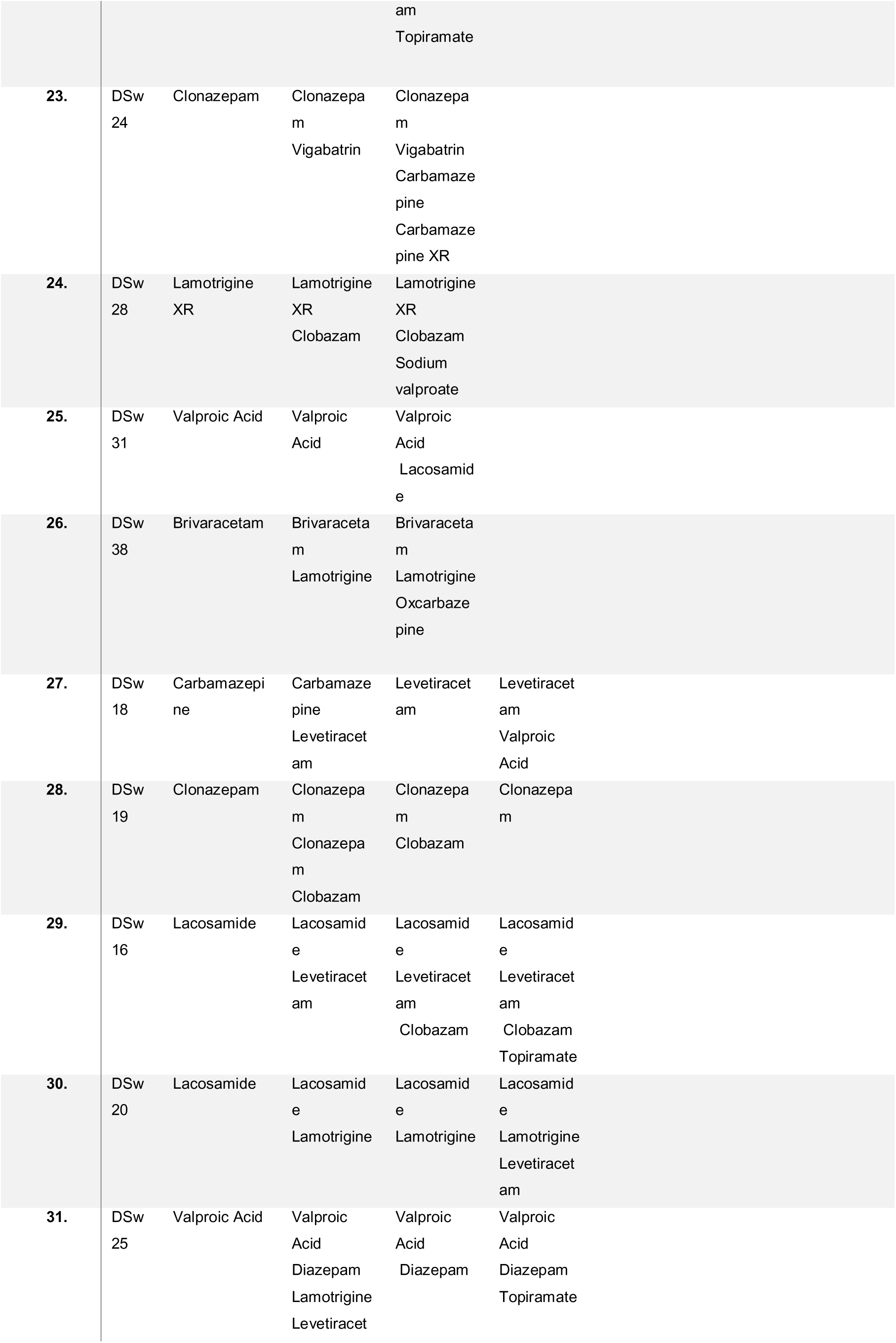
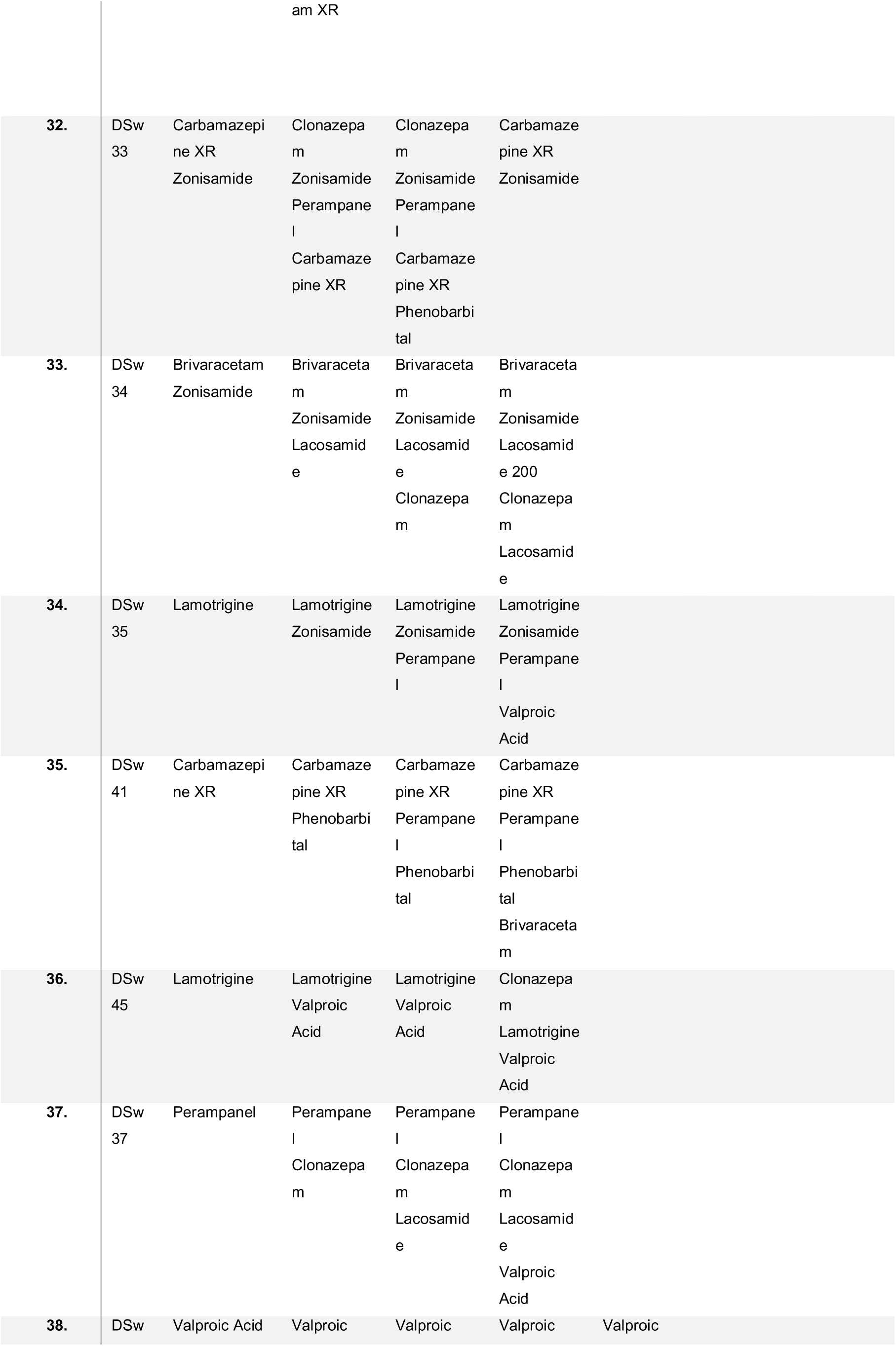
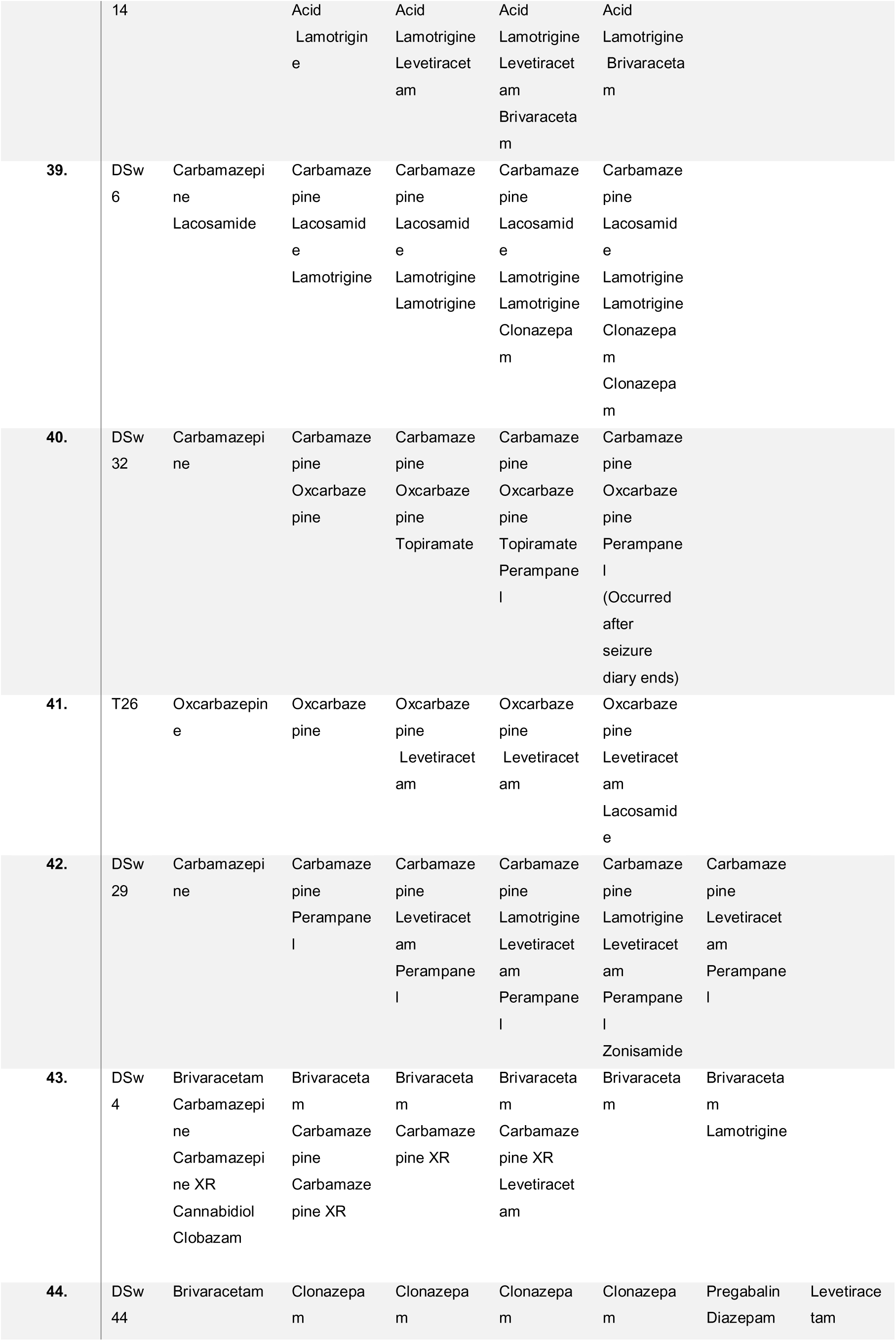
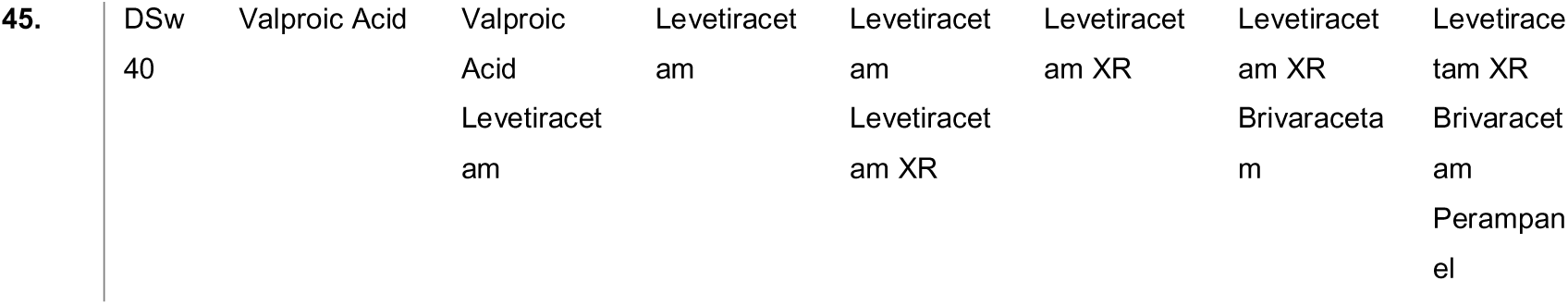
List of all participants, their initial antiseizure medications, and subsequent medication changes. DSu: drug-sustained, DSw: drug-switching.

**Supplementary Table 3.** The frequency of antiseizure medication (ASM) changes.

| FREQUENCY OF PARTICIPANTS WHO HAD ANTISEIZURE MEDICATION CHANGES | DRUG-SUSTAINED<br>(n = 41) | DRUG-SWITCHING<br>(n = 45) |
| --- | --- | --- |
| No drug change | 41 | n/a |
| 1 drug change | N/A | 12 |
| 2 drug changes | N/A | 11 |
| 3 drug changes | N/A | 8 |
| 4 drug changes | N/A | 10 |
| 5 drug changes | N/A | 2 |
| 6 drug changes | N/A | 1 |
| 7 drug changes | N/A | 1 |

**Supplementary Table 4.** The frequency of participants on antiseizure medication (ASM) mono-or polytherapy. There was no statistically significant difference between drug-sustained and drug-switching groups Χ^2^ = 6.00, p=0.54.

| FREQUENCY OF PARTICIPANTS ON MONO- OR POLYTHERAPY | DRUG-SUSTAINED<br>(n = 41) | DRUG-SWITCHING<br>(n = 45) |
| --- | --- | --- |
| No ASM | 0 | 0 |
| 1 ASM | 23 | 32 |
| 2 ASMs | 9 | 7 |
| 3 ASMs | 6 | 3 |
| 4 ASMs | 1 | 2 |
| 5 ASMs | 2 | 1 |

**Supplementary Table 5.** The frequency of participants using antiseizure medications (ASMs) by drug class. There was no significant difference between the drug-sustained group and the drug-switching groups, Χ^2^ = 70.13 p = 0.10.

| FREQUENCY OF PARTICIPANTS USING<br>ASM DRUG CLASSES | DRUG-SUSTAINED<br>(n = 41) | DRUG-<br>SWITCHING<br>(n = 45) |
| --- | --- | --- |
| AMPA-glutamate receptor inhibitor | 5 | 6 |
| Carbonic anhydrase inhibitor | 1 | 0 |
| GABA<sub>A</sub> receptor agonist | 18 | 20 |
| GABA transaminase enzyme inhibitor | 0 | 1 |
| Modulation of alpha-2 delta subunit of<br>voltage-gated calcium channels | 4 | 2 |
| Modulation of synaptic vesicle protein 2A | 15 | 28 |
| NMDA-glutamate receptor inhibitor | 0 | 10 |
| Opening KCNQ2-5 (Kv7.2-Kv7.5) voltage-gated potassium channels | 0 | 0 |
| Type t voltage gated calcium channel<br>inhibitor | 0 | 1 |
| Various actions on multiple targets | 14 | 34 |
| Voltage gated sodium channel inhibitor | 27 | 71 |

**Supplementary Table 6.**
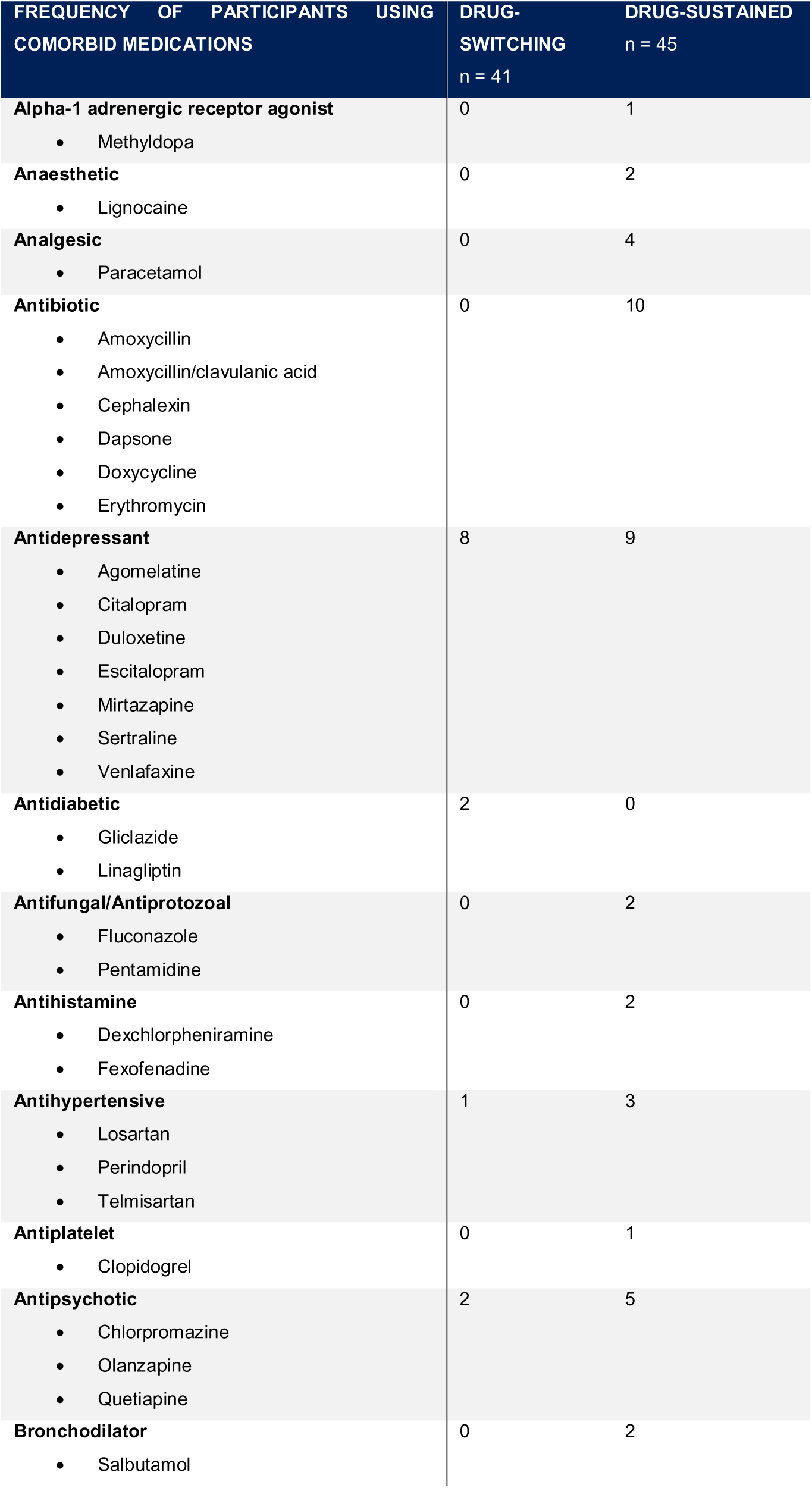
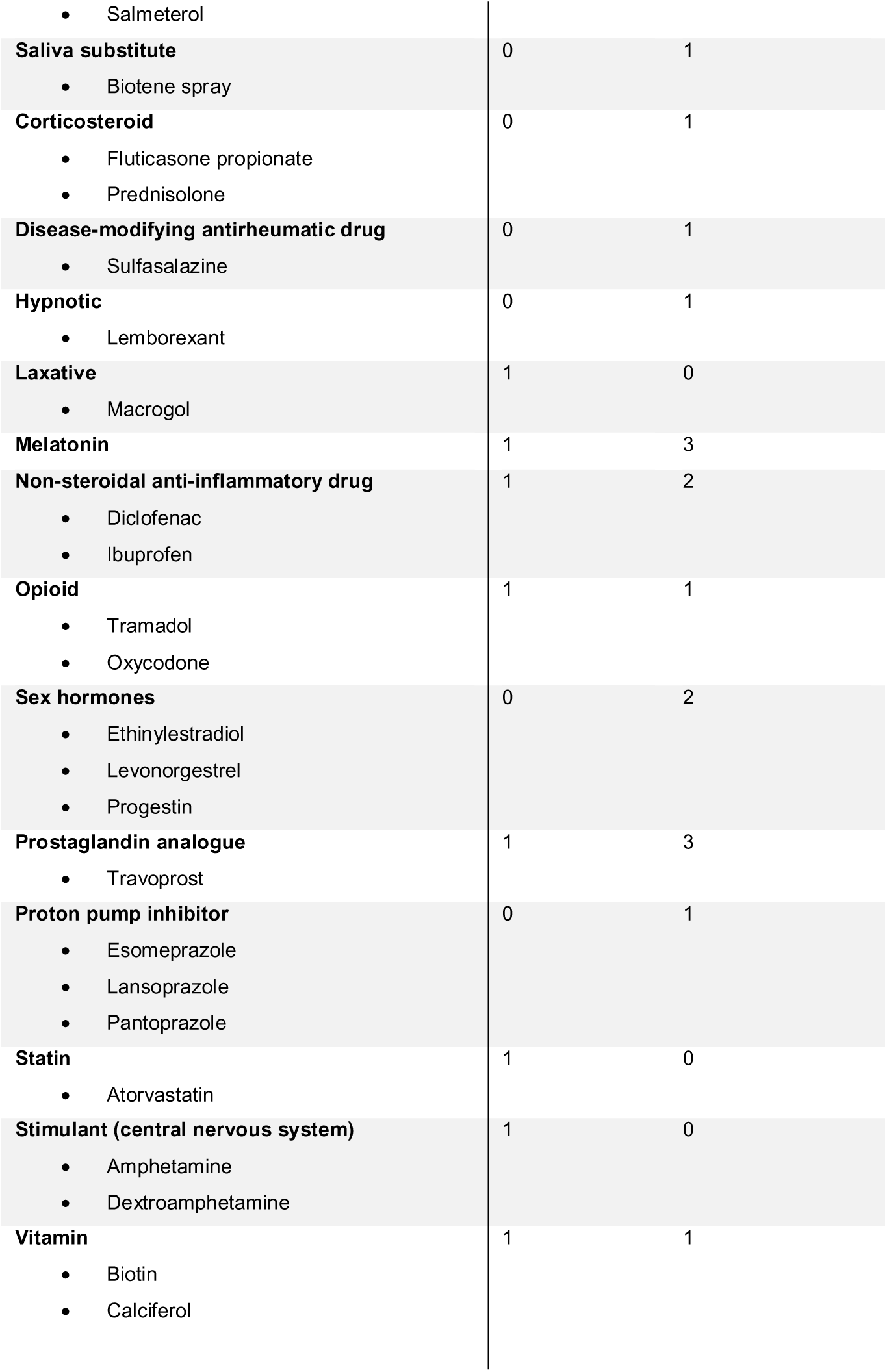
The frequency of participants using comorbid medications by drug class. There was a statistically significant difference between the drug-switching group and drug-sustained group, Χ^2^ = 54.69, p = 7.84e^-5^.

**Supplementary Table 7.**
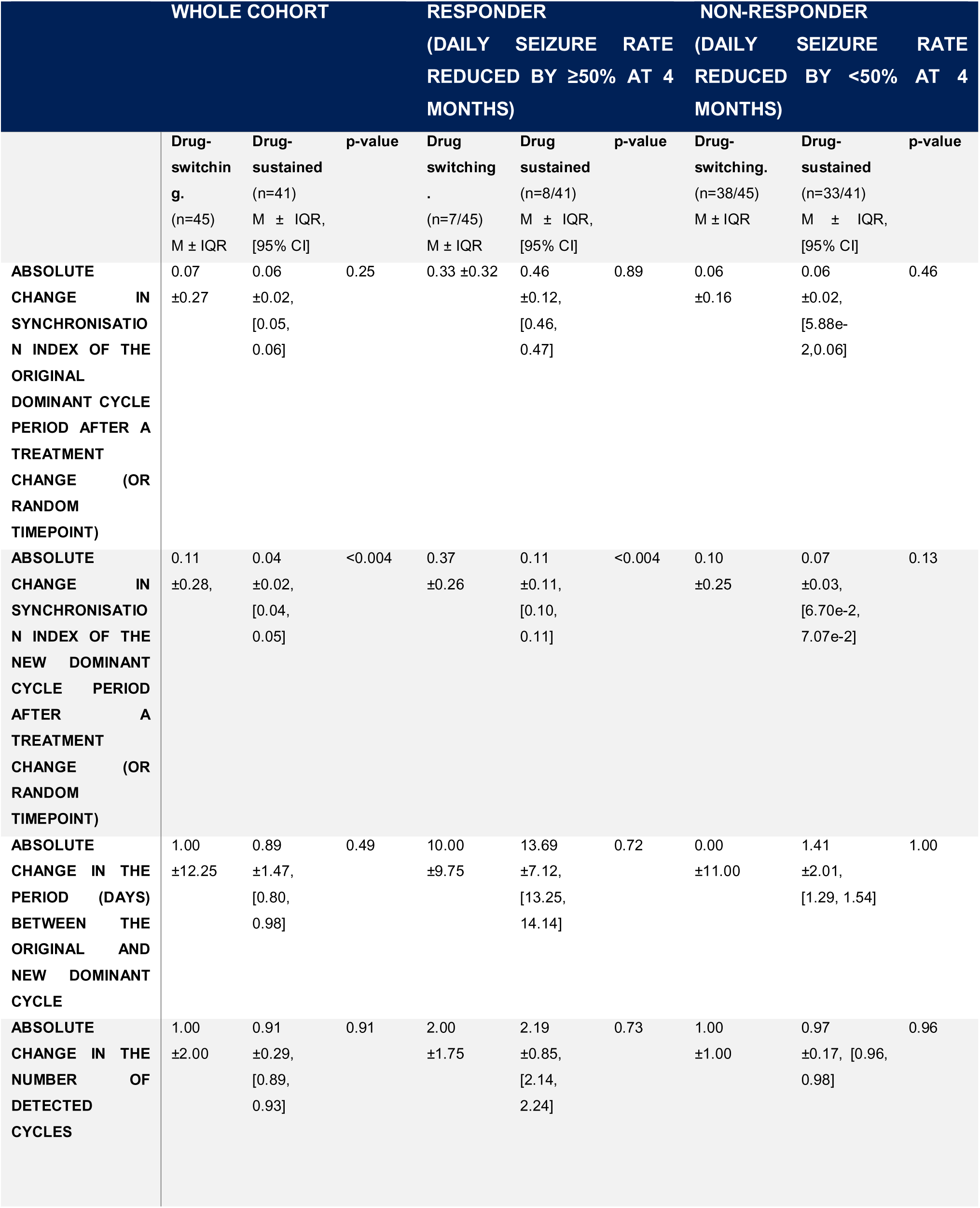
Permutation test results: absolute change in seizure cycle properties after a change in antiseizure medication. The new dominant seizure cycle Synchronisation Index significantly changed when ASMs were altered. This effect is observed in the entire drug-switching group and among responders, but not in non-responders. No other significant differences in seizure cycle properties were identified. M: median, IQR: interquartile range, CI: confidence interval.

**Supplementary Figure 2.**
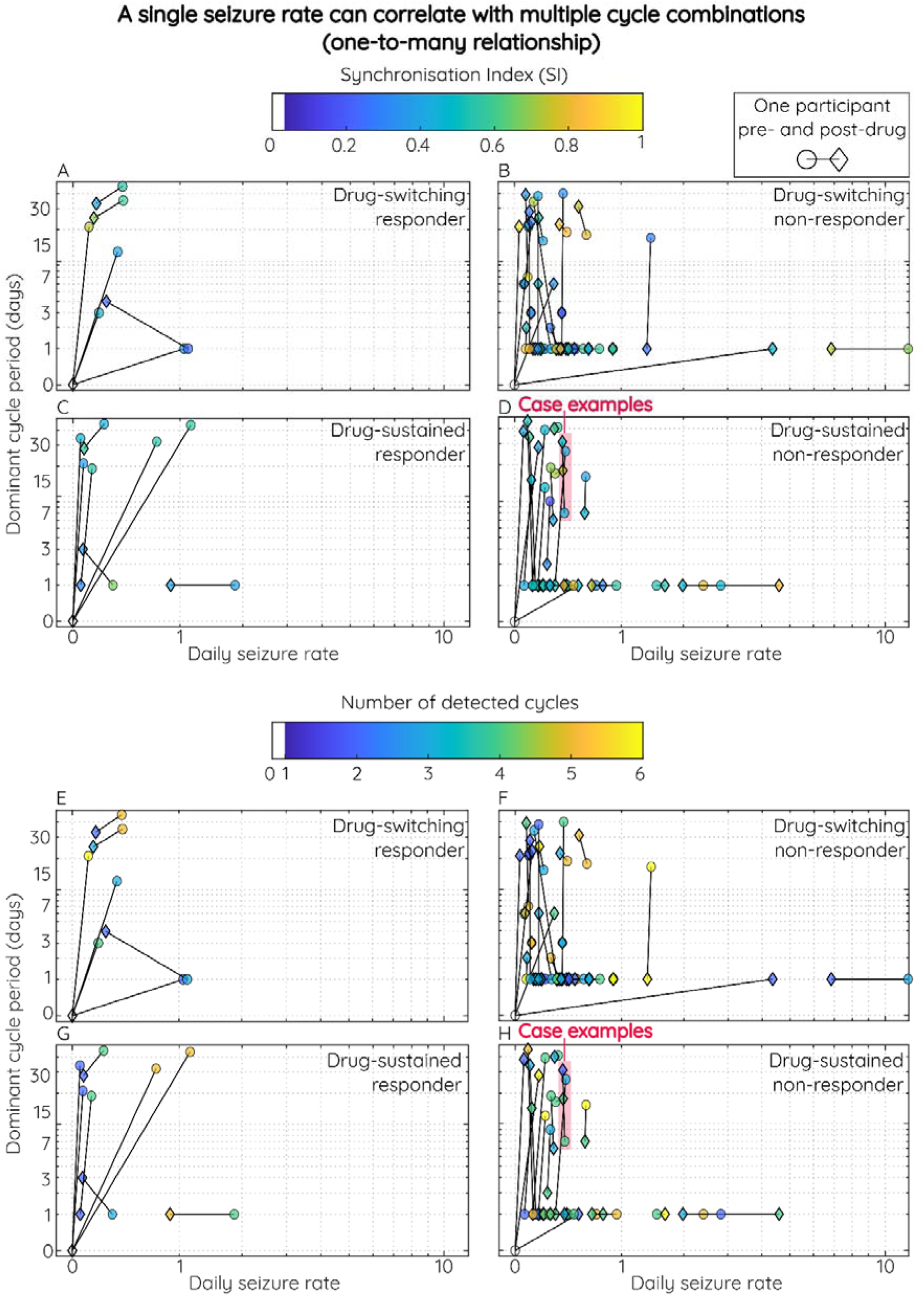
Seizure rates versus seizure cycle properties. (A-D) Similar seizure rates can be associated with different seizure cycle periods with different Synchronisation Index (SI) values, regardless of treatment type or response. This is illustrated by different symbols vertically aligned on the x-axis (daily seizure rate averaged over 4-months). The different symbols include different shapes (circles representing pre-drug or diamond representing post-drug), y-axis positions (indicating the dominant cycle period), and colours. (A-D) Colours show the SI of the dominant cycle. However, in plots (E-H), colours represent the number of detected seizure cycles. Note, the x- and y-axes are logarithmically scaled. A one-to-many relationship may also be observed as similar seizure rates are associated with a different number of detected seizure cycles with different periods. Figure 6D highlights how distinct seizure cycle patterns can emerge despite similar seizure rates, as illustrated by three non-responder drug-sustained participants within the pink rectangular box labelled ‘case examples’. Starting from the bottom of the pink box, Case 1 (pre-drug): represented by a light blue circle, this participant’s seizures synchronise (SI = 0.42) with a 7-day cycle (y-axis) and occur at a rate of ∼46 seizures in 4 months (0.38 seizures/day on the x-axis). Case 2 (post-drug): shown as a green-yellow diamond, their seizures synchronise more strongly (SI = 0.74) with a 17-day cycle (y-axis) and have a similar rate of ∼45 seizures in 4 months (0.37 seizures/day on the x-axis). Case 3 (pre-drug): represented by another light blue circle, this participant’s seizures synchronise (SI = 0.40) with a 25-day cycle and occur at a rate of ∼46 seizures in 4 months (0.38 seizures/day on the x-axis). Case 1 (post-drug): shown as a green-blue diamond at the top of the pink box, this participant’s seizures now synchronise more strongly (SI = 0.48) with a 30-day cycle, while they had ∼45 seizures in 4 months (0.37 seizures/day on the x-axis). Figure 6H, highlights the same case examples with similar seizure rates 0.37-0.38 seizures/day, but the colour axis indicates the number of detected seizure cycles: Case 1 pre-drug has 3 cycles (green circle), Case 2 pre-drug also had 3 cycles (green circle), Case 3 pre-drug had 2 cycles (light blue circle), and Case 1 post-drug only had 1 cycle detected (dark blue diamond). The observed vertical alignment on the x-axes of these three case examples, as well as the vertical alignment of other participants within Figure 3, illustrates that similar seizure rates can correspond to a wide range of cycle variables, including strength, duration, and the number of detected cycles. This provides preliminary evidence to suggest a one-to-many relationship exists between seizure rate and seizure cycle variables.

